# Genome-by-Trauma Exposure Interaction Effects in Depression

**DOI:** 10.1101/2022.03.11.22272206

**Authors:** T.M. Chuong, M.J. Adams, A.S.F. Kwong, C.S. Haley, C Amador, A.M McIntosh

## Abstract

**Background:** Self-reported trauma exposure has consistently been found to be a risk factor for Major Depressive Disorder (MDD) and several studies have reported interactions with genetic liability. To date, most studies have examined interaction effects with trauma exposure using genome-wide variants (single nucleotide polymorphisms SNPs) or polygenic scores, both typically capturing less than 3% of phenotypic risk variance. We sought to re-examine genome-by-trauma interaction effects using genetic measures utilising all available genotyped data and thus, maximising accounted variance.

**Methods:** Measures of self-reported depression, neuroticism and trauma exposure for 148 129 participants with whole genome SNP data are available from the UK Biobank study. Here, we used a mixed-model statistical approach utilising genetic, trauma exposure and genome-by-trauma exposure interaction similarity matrices to explore sources of variation in depression and neuroticism.

**Findings:** Our approach estimated the heritability of MDD to be approximately 0·160 [SE 0·016]. Subtypes of self-reported trauma exposure (catastrophic, adult, childhood and full trauma) accounted for a significant proportion of the variance of each trait, ranging from 0·056 [SE 0·013] to 0·176 [SE 0·025]. The proportion of MDD risk variance accounted for by significant genome-by-trauma interaction ranged from 0·074 [SE 0·006] to 0·201 [SE 0·009]. Results from sex-specific analyses found genome-by-trauma interaction variance estimates approximately 5-fold greater for MDD in males than in females.

**Interpretation:** This is the first study to utilise an approach combining all genome-wide SNP data when exploring genome-by-trauma interaction effects in MDD and present evidence that interaction effects are influential in depression manifestation. This effect accounts for greater trait variance within males which points to potential differences in depression aetiology between the sexes. The methodology utilised in this study can be extrapolated to other environmental factors to identify modifiable risk environments and at-risk groups to target with interventions.

**Research In Context:** *Evidence before this study:* We searched PubMed up to January 30^th^ 2022, with the following terms: (“gene environment interaction” OR “gene environment” OR “genome wide by environment” OR “GWEIS” OR “polygenic environment” OR (“gene” AND “environment”)) AND (“polygenic risk score” OR “polygenic score” OR “genomic relationship matrix” OR “GRM”) AND (“trauma” OR “environmental adversity” OR “stressful life events”) AND (“depression” OR “major depressive disorder” OR “MDD” OR “depressive symptoms”). Date or language restrictions were not applied. We further reviewed the reference lists of identified articles. This search was supplemented by reviewing related articles identified by Google Scholar. We identified 12 relevant articles. Studies to date have not explored genome-by-environment interaction effects in depression using genomic similarity matrices, however, these effects have been explored using individual single nucleotide polymorphisms (SNPs) from genome-wide studies and polygenic scores (PGSs). Some findings suggest genome-by-environment interaction effects increase risk of depression. However, replication attempts have produced either inconsistent or null findings. Taken together, it is evident that findings have failed to provide consistent evidence of substantial interaction effects. These findings may be a result of limited statistical power in analyses due to genome-wide variants and PGSs failing to capture the polygenic nature of depression with sufficient precision.

*Added value of this study:* This study is the first to explore genome-by-trauma interaction effects on MDD through the estimation of variance components using relationship matrices. Genomic relationship matrices (GRMs) utilise all available genotyped variants, thus, capturing a greater proportion of the trait variance and potentially providing greater power to detect genetic effects in comparison to PGSs. Additional relationship matrices capturing trauma exposure, and genome-by-trauma exposure similarity are computed and included into mixed linear models. We found evidence for substantial genome-by-trauma (including subtypes of trauma) exposure interaction effects on depression manifestation. Estimated genome-by-trauma interaction effects were larger in males than in females.

*Implications of all the available evidence:* Our findings are the first to show substantial genome-by-trauma effects on depression using whole genome methods. These findings highlight that the role of trauma exposure on depression manifestation may be non-additive and different between sexes. Exploring these effects in depth may yield important insight into various mechanisms, which may explain prevalence differences observed between males and females. Future work can build upon the framework we propose to explore genome-by-trauma interaction effects and the underlying molecular sites and mechanisms which are involved in depression manifestation.

## Introduction

Depression is a highly prevalent psychiatric disorder with a lifetime risk of ∼16%^1, 2^ and is a leading cause of disability worldwide.^3^ Twin studies have produced heritability estimates ranging between 30-40%^4^ suggesting both genetic and environmental factors are influential. In fact, genome-wide association studies (GWAS) have uncovered many single nucleotide polymorphisms (SNPs) associated with depression, however, SNP-based heritability estimates of 5-10%^5, 6^ are much lower than estimates obtained from twin studies which has highlighted the issue of missing heritability. Explanations for missing heritability include inflation of twin-based heritability estimates attributable to shared environmental effects as well as gene-environment interplay.^7-10^

Some environmental factors such as self-reported trauma exposure have been found to play a role in depression; with case-control studies suggesting individuals diagnosed with Major Depressive Disorder (MDD) report higher levels of trauma exposure.^11-13^ In turn, trauma exposure in childhood has been consistently associated with adverse outcomes including increased risk of MDD in adolescence and adulthood.^14, 15^ These findings strongly suggest a causal role for trauma in depression’s aetiology. Moreover, research has shown self-reported trauma exposure and MDD to be genetically correlated, suggesting shared genetic risk factors for both^16^, or potentially more complex interplay effects of genetics and trauma exposure on depression manifestation.

One key form of interplay that has been explored is the interaction effect of genetics and trauma exposure on depression manifestation. Gene-by-environment interaction effects refer to the differential effects of an environmental exposure on traits in individuals with differing genotypes. This can be conceptualised as an individual’s genetic sensitivity to certain environments, which may result in an exacerbated risk of a disorder.^17, 18^ Minimal evidence of interaction effects has been yielded from studies exploring SNP-by-trauma effects.^16, 19-22^

Moreover, research utilising polygenic scores (PGSs); genetic measures that can be calculated for each individual by identifying, weighting, and summing genotyped risk variants found to be associated with depression^23, 24^, have yielded inconsistent findings. Whilst some studies have highlighted sex differences and found significant interaction effects associated with MDD outcomes^18, 25-27^, some replication attempts reported null findings^28, 29^ and follow up meta-analyses have suggested that reported findings were likely to be false positives.^30^ An explanation for the inconsistent findings may lie in the predictive accuracy and validity of PGSs.^10^ PGSs build upon the information provided by GWAS, which still have limited statistical power for detecting trait associated genetic variants and their effect sizes.^24, 31, 32^ Power limitations of GWAS, and thus PGSs, are greater for traits that also have a substantial environmental component, such as depression, as opposed to traits with higher genetic aetiology.^33^ This is reflected in the fact that current PGSs capture less than 3%^5, 6^ of the phenotypic variance proposed for depression.

One way to circumvent the issues associated with PGSs, is to make use of genetic measures that are capturing greater variance. Genomic relationship matrices (GRMs) have been able to do this by utilising all genotyped SNPs.^34^ Including matrices representing genetic, environmental and interaction effect similarity within a sample, in mixed linear models, can provide estimates of the genetic, environmental, and genome-by-environment interaction components of trait variance.^35^

Here, we estimated the contribution of trauma exposure and its interaction with genetic variation to depression as well as neuroticism. We chose to explore neuroticism as this trait has been shown to have a greater genetic component^36, 37^ and has a strong phenotypic link with MDD suggesting the exploration of neuroticism to be useful in understanding the genetic aetiology of both of these traits.^38, 39^ As the existing literature has highlighted the role of trauma exposure in sex differences observed in MDD, we also explored these effects in males and females separately.^40^ Here we show that utilising the entirety of genotyped genetic variants can improve statistical power in the exploration of genome-by-trauma interaction effects. More importantly, our findings suggest that genome-by-trauma interaction effects may play a much larger role in depression manifestation than previously thought.

## Methods

### PARTICIPANTS

We used data from the UK Biobank (UKB), a national study exploring genetic and environmental determinants of health using individuals recruited from 22 different centres across the United Kingdom.^41, 42^ A follow up Mental Health Questionnaire (MHQ) was administered assessing common mental health disorders including trauma experience.^43^ In this study we had access to individual level data from 148 129 participants who completed the MHQ with available genetic, trauma experience, depressive symptoms and/or neuroticism information.

The UKB study received ethical approval from the NHS National Research Ethics Service (reference: 11/NW/0382) and all participants provided written informed consent. This study has been approved by the UKB Access Committee (Project #4844).

### GENOTYPES

Two separate array chips with 95% SNP overlap, were used to genotype UKB participants; the Applied Biosystems™ UK BiLEVE Axiom™ Array by Affymetrix1 captures 807 411 SNPs and the Applied Biosystems™ UK Biobank Axiom™ Array captures 825 927 SNPs.^44^ Quality control of genotyped SNPs consisted of the exclusion of SNPs with missingness > 2% and a Hardy-Weinberg Equilibrium test p < 10^−6^. SNPs with minor allele frequency (MAF) < 0.05 and individuals with > 2% missing genotypes were excluded from analyses. A total of 414 584 common SNPs across the 22 autosomes were included in analyses.

### PHENOTYPES

All phenotypes were defined using retrospective self-reported responses to questions assessing help-seeking behaviour, depressive symptoms, and trauma experience. Field codes and more information on classifications are available in supplementary table 1.

#### Depression & Neuroticism Phenotypes

CIDI depression was defined using questions from the Composite International Diagnostic Interview Short Form (CIDI).^45^ These were administered within the online follow-up Mental Health Questionnaire (MHQ), directly assessing the Diagnostic and Statistical Manual of Mental Disorders, Fifth Edition (DSM-5)^46^ symptomatic and functional criteria for MDD, and are associated with greater MDD specificity.^47^

Supplementary analyses were conducted using broad depression and neuroticism phenotypes. Broad depression was defined using self-reported help seeking behaviour for mental health difficulties. Case and control status was determined from the response to Touchscreen Questionnaire questions administered during initial recruitment; ‘Have you ever seen a general practitioner (GP) for nerves, anxiety, tension or depression?’ or ‘Have you ever seen a psychiatrist for nerves, anxiety, tension or depression? Individuals responding ‘Yes’ to either and ‘No’ to both questions were classified as cases and controls, respectively.

Neuroticism was measured using the 12-item Neuroticism scale of the revised short form Eysenck Personality Questionnaire (EPQ-R).^48^ Response options were ‘Yes’, ‘No’, ‘Do not know’, ‘Prefer not to answer’. Participant summed scores of ‘Yes’ responses (ranging from 0-12) were used in downstream analyses.

#### Trauma Phenotypes

Participants were administered a 16-item questionnaire relating to traumatic experiences as a part of the MHQ. Five items explored childhood trauma using the Childhood Trauma Screener^49, 50^; five items explored adult trauma using an equivalent screener developed by the UKB Mental Health steering group^51^; and the remaining six items explored catastrophic trauma using questions related to events that often trigger post-traumatic stress disorder.^16^ More information on questions, distribution of responses can be found in supplementary figure 1 and table 2. **Table 1** shows samples sizes of individuals included in statistical analyses with complete trauma exposure, depression and neuroticism information.

**Table 1.**
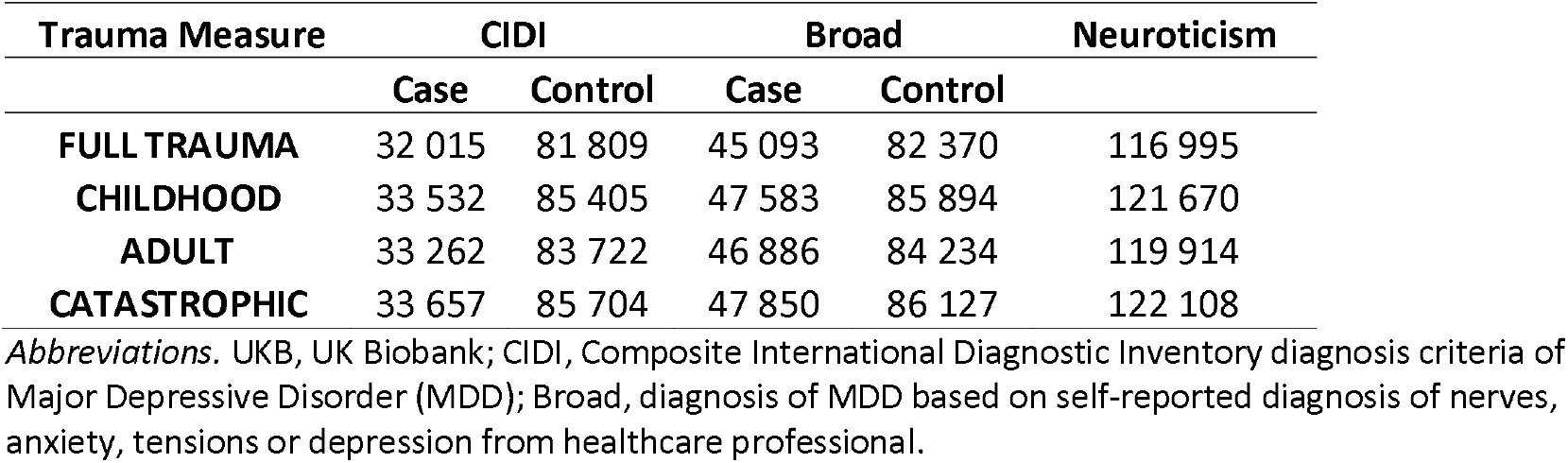
Sample sizes of UKB participants with complete trauma, depression and neuroticism information.

**Table 2.**
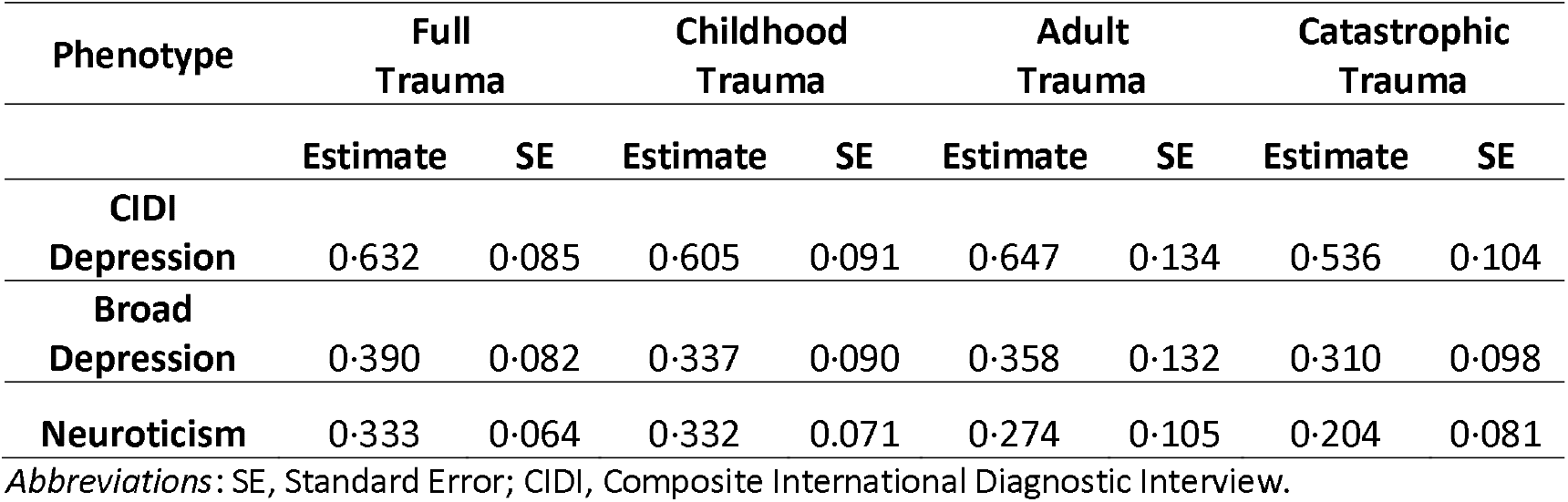
Genetic Correlations Between Trauma and Depression/Neuroticism Phenotypes

Individual regression analyses exploring associations between each trauma item and depression/neuroticism outcomes with age and sex as covariates (supplementary table 2), suggested most trauma items to be significantly associated with all of the three phenotypes. Hence, all items were utilised in the formation of trauma phenotypes.

In order to capture separate and independent dimensions of trauma, principal components (PCs) of complete responses to all trauma items (full trauma) as well as the three sub-categories of trauma items (childhood, adult, catastrophic trauma) were obtained. Trauma items were rescaled to have a mean of one and unit variance before PCS were extracted using the ‘prcomp’ function in R 4.0.2.^52^ For more information on depression/neuroticism associations with trauma PCs and PC loadings see supplementary table 3-4.

### COVARIANCE MATRICES

Covariance matrices were used in the exploration of genetic, environmental and genome-by-environment components of variation. As the number of participants with available depression/neuroticism and trauma information is large (∼150k), analysing the pairwise covariance matrices jointly for the whole cohort was computationally intractable. To work around these computational issues, participants were split into five different clusters based on the geographical location of their recruitment centres (north, midnorth, midsouth, southwest and southeast regions in the UK) and all subsequent analyses were replicated across the five clusters; for more information on how clusters were formed and their demographic characteristics see supplementary Figure 2 and table 5-6.

Genomic Relationship Matrices (GRMs): represent the expected genetic similarity between individuals. We computed one GRM for all individuals in each of the geographical clusters using genetic variants that passed quality control (see section ‘GENOTYPES’) using GCTA v1.91.4beta.^34^ Supplementary GRMs were computed using only unrelated individuals (individuals with genomic similarity values < 0.05).

Trauma Environmental Relationship Matrices (E): represents similarity between individuals based on their trauma eigenvectors (PCs). Separate Es were computed by calculating participant similarity based on their full trauma, childhood, adult and catastrophic trauma eigenvectors. Supplementary Es were computed using the first eigenvector (PC1) of full trauma, as this accounted for the greatest variance in our outcome phenotypes. To control for genetic covariance between trauma exposure and our phenotypes of interest (depression/neuroticism), supplementary Es were computed using trauma eigenvectors pre-corrected for the full GRM. We used OSCA (v0.45) default algorithm 1^53^ to compute these variables, for more information on available algorithms see supplementary materials section A.

Gene-Environment Interaction Matrices (GxE): represent shared genome-by-trauma interaction effects. These were computed by multiplying GRM and ERMs using a cell-by-cell (Hadamard) product.35, 54, 55

### ANALYSES

#### Genetic Correlations

Using the first eigenvector (PC1) for the subtypes of trauma, the SNP heritability of each trauma variable was explored. Heritability estimates of the trauma variables were obtained by fitting the trauma variables as the dependent variable and Gs as random effects within a mixed linear model framework (estimates of trauma PC1 variance attributable to G), see model 1 below. The genetic correlations between trauma PC1 and depression variables were explored using the moment-based method, Haseman-Elston regression analyses, where standard errors were calculated using a leave-one-individual-out jackknife technique. Age, sex, genotyping array and the first 15 principal components of the GRM were included as covariates.

#### Variance Components Analyses

Variance components of depression/neuroticism were explored within mixed linear model frameworks. Four models were explored with varying levels of complexity:

1. *y* = *Xβ* + *g* + *ε*
2. *y* = *Xβ* + *e* + *ε*
3. *y* = *Xβ* + *g* + *e* + *ε*
4. *y* = *Xβ* + *g* + *e* + *g* × *e* + *ε*

Where *y* is a *n* × 1 vector of observed depression/neuroticism phenotypes; *β* is a vector of fixed effects (which include age, sex, genotyping array and the first 15 principal components of the full sample GRM) and *X* is its design matrix; *g* is a *n* × 1 vector of SNP effects (representing additive genetic effects) with 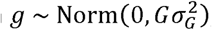; *e* is a *n* × 1 vector representing common environmental effects of childhood, adult, catastrophic or all trauma with 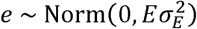; *g* × *e* is a *n* × 1 vector representing interactions between genetic and trauma effects with 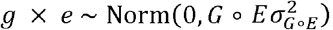; and *ε* is a *n* × 1 vector of residual effects.

Estimates of variance attributable to the G, E and GxE components are obtained from analyses using CIDI and Broad depression as dependent variables and are converted to the liability scale within GCTA.^34^ As UKB is a population-based study, we utilised the prevalence rates observed within the whole sample. Prevalence rates used were 0·28 and 0·35 for joint sex analyses; 0·35 and 0·43 for female analyses; 0·19 and 0·27 for male analyses for CIDI and broad depression, respectively.

Analyses were repeated using only unrelated individuals. All analyses were replicated across the five geographical cluster samples using GCTA v1.91.4beta^34^ and results (estimates of variance components) were meta-analysed using R package ‘metafor’.

## Results

Initial analyses explored the SNP heritability of the first PC of full trauma and sub-categories of trauma. Trauma variables meta-analysed SNP heritability estimates were; full trauma 0·17 [SE 0·008], childhood trauma 0·15 [SE 0·008], adult trauma 0·063 [SE 0·008] and catastrophic trauma 0·11 [SE 0·008] (see supplementary tables 8). Similar results were obtained when using only unrelated individuals. All genetic correlations between the first PC of trauma variables and depression/neuroticism phenotypes are shown in Table 2. Genetic correlations between the first PC of trauma variables and broad depression/neuroticism phenotypes were modest; in contrast, we observed stronger genetic correlations between trauma variables and the CIDI depression phenotype. Results from each cluster can be found in supplementary table 9.

**Figure 1** shows the estimates for the proportion of CIDI depression variance explained by the different sources included in the mixed linear models (results are the meta-analysis of the five UKB sub-samples). All estimates for proportion of variance explained by all components were statistically significant. Log-likelihood ratio tests (LRTs) suggested that the inclusion of trauma (E) and genome-by-trauma (GxE) interaction components improve model fit. Full details of these analyses, including estimates, standard errors (SEs), LRT values as well as results using broad depression and neuroticism as the dependent variable can be found in supplementary materials (supplementary tables 10-13).

**Figure 1.**
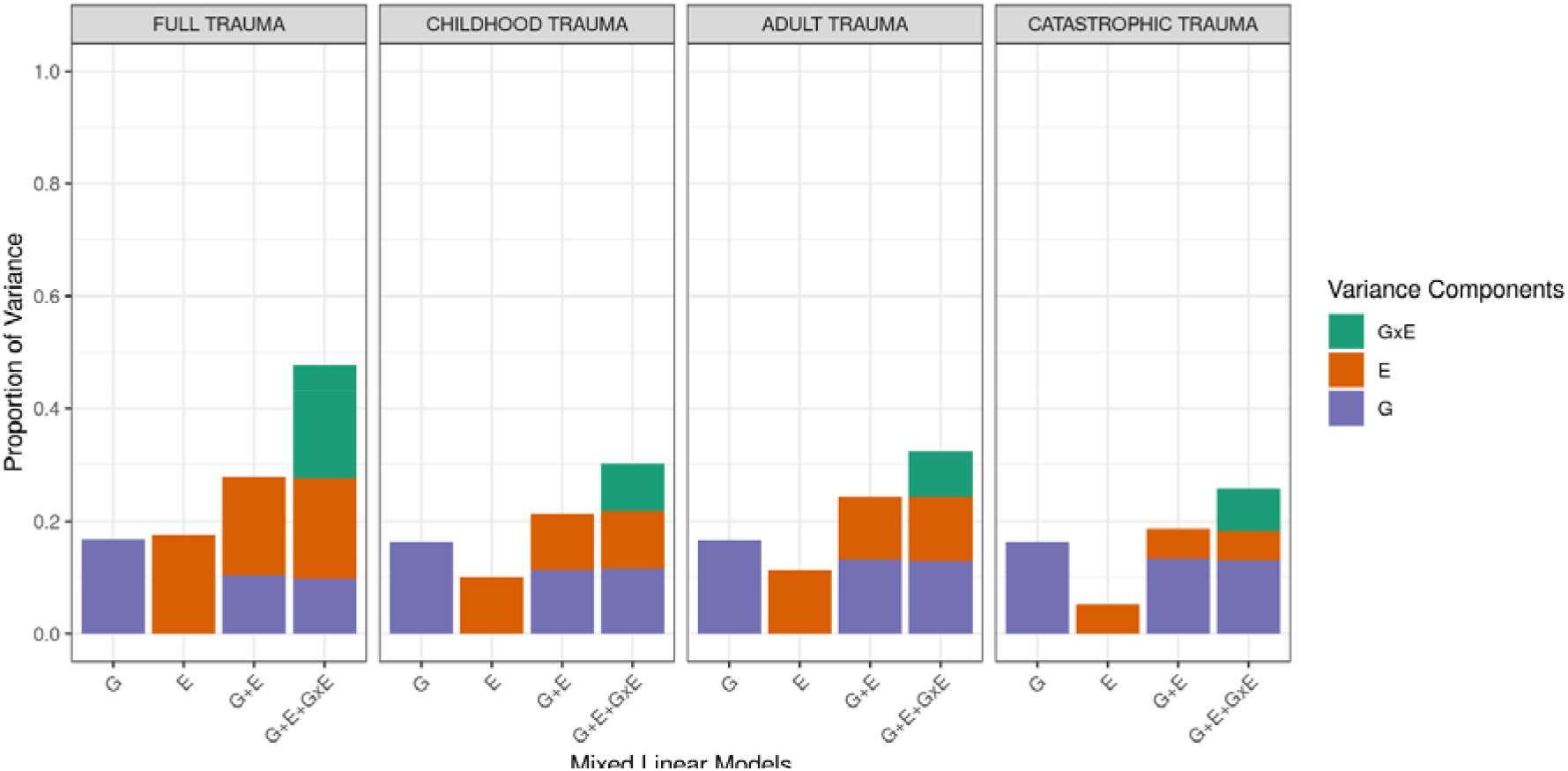
Proportion of CIDI Depression Variance Explained by Genetic, Environmental and Interaction Sources in UK Biobank. The proportion of variance explained is specified on the y-axis. The sources of variation: G, Genomic; E, Environmental; GxE, Gene-by-Environment Interactions and their corresponding models are specified on the x-axis. E represent trauma exposure. Facets show the different trauma exposures explored.

Heritability estimates (i.e. proportion of phenotypic variance accounted for by the G) of CIDI depression were stable across the different UKB sub-samples ∼ 0·16 [SE 0·016]. The meta-analysed estimates for proportion of variance attributable to the trauma relationship matrices (Es) were 0·18 [SE 0·025]=full trauma, 0·101 [SE 0·027]=childhood trauma, 0·113 [SE 0·03]=adult trauma, 0·05 [SE 0·013]=catastrophic trauma. The meta-analyses estimates for proportion of variance attributable to the interaction effect (GxE) were highest when exploring full trauma 0·201 [SE 0·009] and were 0·084 [SE 0·006], 0·081 [SE 0·005], 0·074 [SE 0.006] when respectively exploring childhood, adult and catastrophic trauma separately.

Similar results were obtained when using only unrelated individuals as well as when mixed linear models utilise Es computed from trauma eigenvectors pre-corrected for the full sample G (supplementary tables 14-15). In contrast, whilst model fit, compared to models excluding Es, are significantly improved, we observed smaller estimates and LRT values when mixed linear models utilised Es computed from only the first eigenvector (PC1) of full trauma items (supplementary table 1).

Significant, yet, smaller estimates of variance components were observed for broad depression and neuroticism (supplementary tables 10-13).

**Figure 2** shows the estimates for the proportion of CIDI depression variance explained by the interaction (GxE) included in the mixed linear models. Here, the interaction effect used Es capturing full trauma exposure. Results for each geographical cluster as well as within female/male only samples are presented. Full details of these analyses, including estimates, SEs, LRTs values as well as results using broad depression and neuroticism as the dependant variable can be found in supplementary materials (supplementary tables 10,17-18).

**Figure 2.**
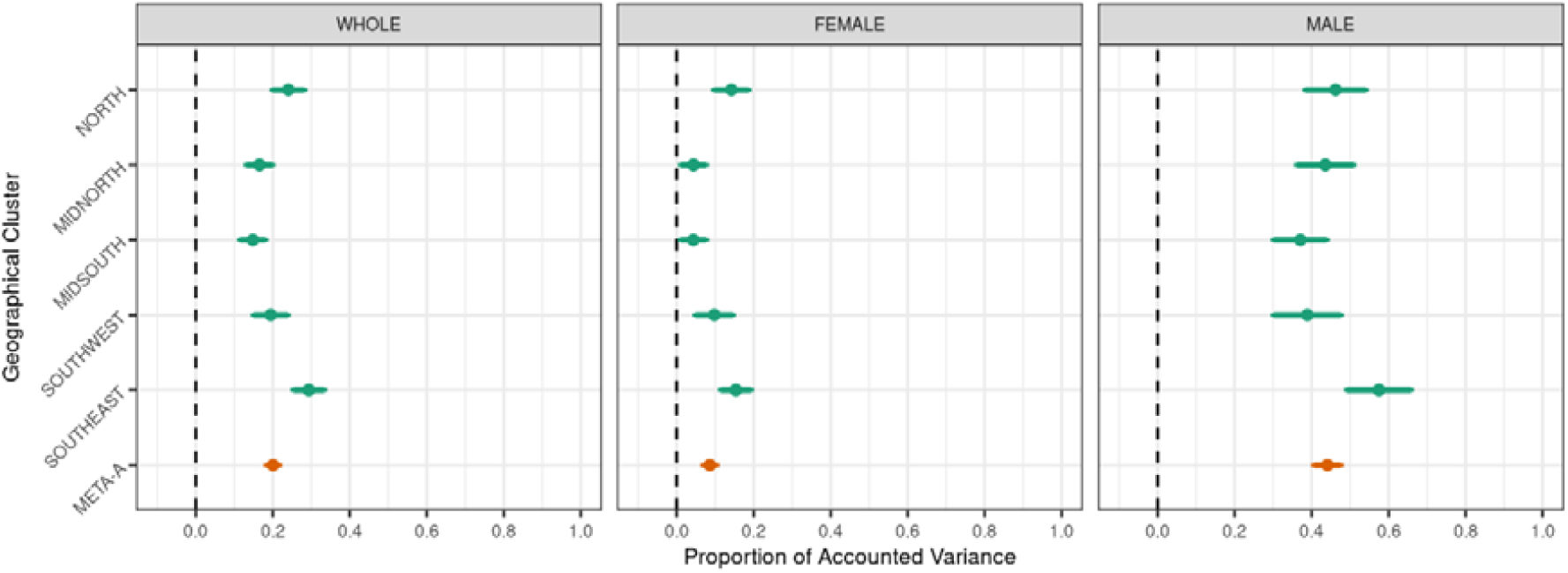
Proportion of CIDI Depression Variation explained by the Genome-by-Trauma Interactions. Forest plot x-axis shows proportion of CIDI Depression variance explained by the genome-by-trauma interaction effect with bars representing standard errors. Results for geographic clusters (green) and meta-analysed estimates (orange) are shown on the y-axis. Facets represent the analysis results using different samples; WHOLE, joint male and female sample; FEMALE, female only sample; MALE, male only sample.

Meta-analysed estimates for proportion of CIDI depression variance explained by the interaction effect across the clusters were statistically significant within the whole (joint female and male), female and male samples. Compared to the analyses of the full sample (joint male and female), the meta-analysed interaction variance was smaller when explored within the female sample and increased when explored within the male sample. Similar results were observed when using only unrelated individuals (supplementary tables 19-20).

## Discussion

This study sought to provide a comprehensive update to the literature on genome-by-trauma interaction effects within depression and depression related traits by exploring variance components as opposed to fixed effect estimates of genome-by-trauma interactions. We compute genetic (using all genotyped variants), trauma exposure and genome-by-trauma interaction effect similarity to explore trait variance attributable to these effects. By utilising all available genotyped data, the mixed linear models implemented have greater statistical power to identify phenotypic variation attributable to genetic and genome-by-trauma interaction effects. We explore general (full trauma) and specific (childhood, adult, catastrophic) measures of trauma.

Our heritability estimates of the trauma measures support findings from the literature.^56^ Our results suggest statistically significant and modest genetic correlations between the trauma and depression/neuroticism variables. Genetic correlations are 2-fold greater with CIDI depression as opposed to broad depression and neuroticism. A perception component of these traits i.e. the more extreme one is on the depression scale the more likely one is to either remember or perceive an event as traumatic, may explain these genetic correlations.

Our results provide depression and neuroticism heritability (proportion of variance captured by the common additive genetic variants) estimates and estimates for proportion of variance attributable to self-reported trauma exposure in line with previous literature.^57-59^ Findings also suggest the sub-categories; childhood, adult and catastrophic trauma explain a substantial proportion of CIDI depression variance.

This study is the first to show contributions of genome-by-trauma interaction effects to depression (and related traits) phenotypic variation that are of relatively large magnitude (7-20%), approximately the same magnitude as the variance captured by self-reported trauma exposure itself (5-18%). These are consistent (although estimates are lower) across the sub-categories (childhood, adult and catastrophic) of trauma exposure (see **Figure 1**). Lower, yet still significant, interaction estimates are observed for broad depression and neuroticism phenotypes, except for the non-significant genome-by-catastrophic trauma interaction estimate within broad depression (supplementary table 13).

Model fit is also significantly improved when mixed linear models utilise environmental relationship matrices (Es) computed using only the first eigenvector (principal component (PC) 1) of trauma items. However, these estimates are substantially attenuated when compared to results from models including Es computed using all PCs of trauma items. This suggests that important trauma exposure and genome-by-trauma interaction effects may be distributed across the different dimensions of self-reported trauma exposure. The inclusion of more self-reported trauma exposure PCs, may additionally capture the differential impact of subtypes of trauma exposure. Our research design and analyses are repeated across five geographical cluster samples. These within-sample replications, while not independent samples, yielded relatively consistent estimates and standard errors, increasing confidence in our results.

Exploring variance components of depression/neuroticism within males and females separately indicates that the proportion of CIDI depression variation captured by genome-by-trauma interaction effects differ substantially between the sexes, with estimates being approximately 5-fold greater in males. This pattern, although estimates were lower, was also observed for broad depression and neuroticism phenotypes. These results, alongside the evident prevalence differences, highlight the need to explore these effects within the sexes separately. Our findings suggest that trauma exposure and sensitivity to trauma exposure accounts for greater variance in depression/neuroticism outcomes for males. Whilst utilising principal components (PCs) enables the use of all trauma exposure variables, it is difficult to interpret directions of associations as higher PC values do not necessarily mean higher levels of trauma exposure. Further research can be conducted to explore the direction of these associations. Exploring individual trauma (e.g. neglect, physical abuse etc.) measures may provide a better understanding of the effect of specific trauma and genome-by-trauma experiences.

Our findings provide evidence of differential effects of trauma exposure dependent on differences in individual genetic liability to depression. Results can highlight both modifiable environments for preventative measures as well as at-risk groups that should be prioritised for these interventions. It is evident that the use of all genotyped variants yield substantial findings when exploring genome-by-trauma exposure interaction effects as opposed to much of the literature making use of PGSs.^16, 20, 26, 30^ It is evident that the method employed here has major advantages over polygenic scores (PGSs), when exploring genome-by-environment interaction effects. However, there are limitations to this study that need to be considered when interpreting results.

Whilst, the GRMs computed utilise all genotyped SNPs, and subsequently account for greater variance than PGSs; discrepancies between twin study heritability and SNP heritability estimates of depression are still apparent. Twin study estimates may be biased upward due to the presence of gene-environment interplay effects, and thus, real heritability estimates are likely to fall between SNP heritability and twin study heritability estimates.^60^ Moreover, results show that our environmental variables, full trauma and the sub-categories of trauma, have moderate heritability estimates and these are genetically correlated with our outcome phenotypes (depression/neuroticism). This highlights that our environmental measures capture both genetic and environmental variances. As genomic relationship matrices are not capturing the entirety of the genetic variance within depression/neuroticism outcomes, the variance captured by the trauma (and sub-trauma) measures may capture residual genetic variance.

Here, to control for genetic covariance between our environmental and outcome variables, we also explored measures of trauma pre-corrected for the available genetic measure (GRMs). The differences in estimates of variance components were negligible. However, similar to the aforementioned limitation, this effect can be more accurately controlled for with an improved genetic measure (e.g. GRM utilising imputed or whole-genome sequenced data). Simulation findings from the literature suggests that making use of imputed or whole-genome sequencing genetic data for GRMs can uncover a further substantial proportion of genetic variance^60^, which would be useful in addressing the limitations outlined above (although would increase time and computational resources needed).

Our trauma exposure, depression and neuroticism variables were measured using retrospective self-report. Furthermore, measures of trauma exposure and CIDI depression were obtained later than measures of broad depression and neuroticism with the follow up UKB mental health questionnaire. This indicates potential measurement error within our variables.^16^ However, it is important to note that self-report is likely the most effective method to explore these phenotypes in studies the size of UKB. Incorporating more objective measures of trauma exposure, e.g. omics measures (DNA methylation) may be able to provide a measure of trauma exposure that is less susceptible to reporting bias and thus, measurement error. For instance, the availability of methylation data has and continues to increase substantially, and can be used as good, and in some cases far better proxy measures of environments as seen with smoking.^35, 61^ Evidence suggests there may be a methylation profile observed in individuals with trauma exposure.^62, 63^ Genome-by-environment interaction effects using methylation data, can then be dissected to explore biological pathways with non-additive effects on outcomes that can be directly targeted. Findings could also further clarify the relationship between genetic liability and trauma exposure.

In conclusion, our findings provide empirical evidence of depression/neuroticism variation attributable to genome-by-trauma interaction effects. We further highlight that the magnitude of these effects are much larger for males in comparison to females. These findings can be further explored to identify both risk groups and modifiable environments/biological pathways which yield greater risk of depression manifestation, which would be useful in personalised/preventative interventions.

## Data Availability

All data produced in the present study are available upon reasonable request to the authors

