## supplementary materials for "Genome-by-Trauma Exposure Interaction Effects in Depression"

**Appendix II. Supplementary Materials for Chapter 3 ‘Genome-by-Trauma Interaction Effects on Depression’**

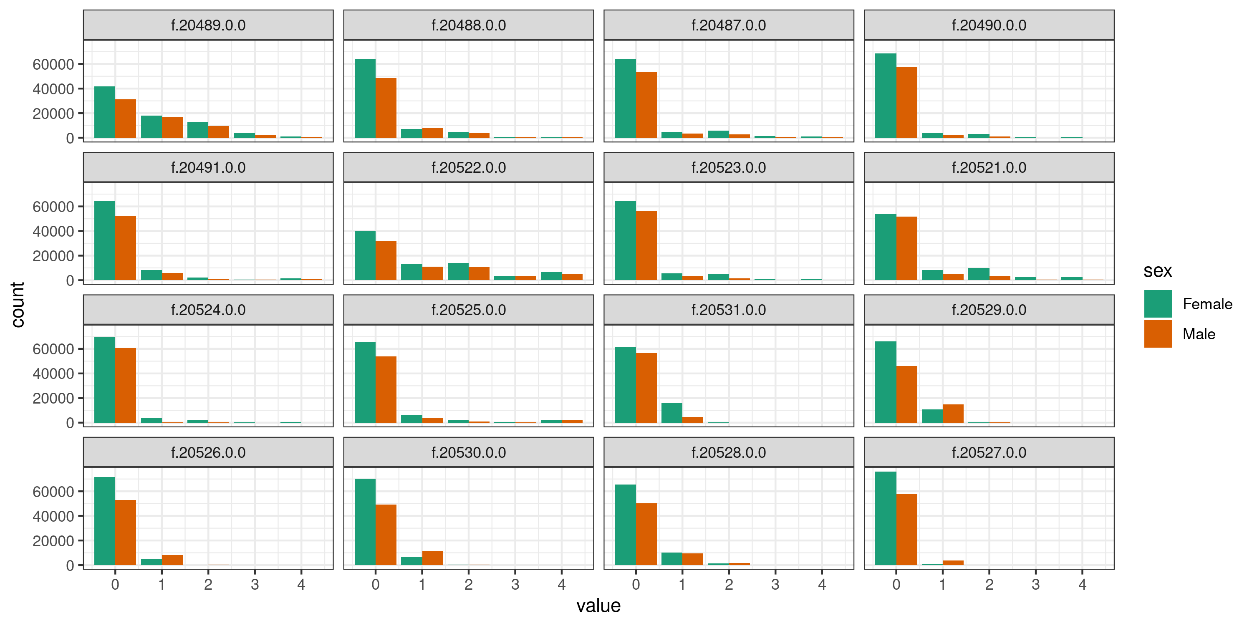

***Figure 1.*** Trauma Exposure Distribution by Sex. Facets represent the 16 different trauma exposure questions (field codes) available within the UK Biobank Mental Health Questionnaire. The x-axis represents the available responses ‘0 - Never True; 1- Rarely True; 2 – Sometimes True; 3 – Often True; 4 – Always True’. The questions related to catastrophic trauma exposure responses were ‘0 – Never True; 1 – Yes, not within 12 months; 2 – Yes, within 12 months’. Each field code and corresponding question can be found in table 2 of this appendix.

**Table 1.1.** Broad Depression UK Biobank Questions and Field Codes

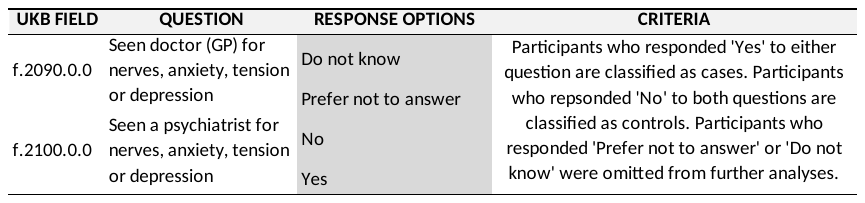

**Table 1.2.** Neuroticism UK Biobank Questions and Field Codes

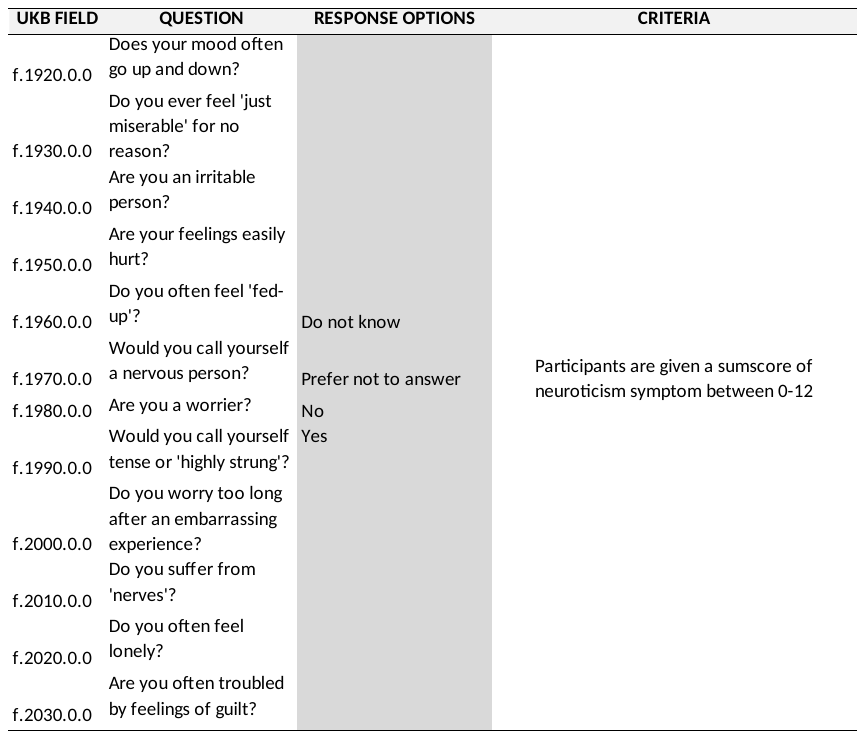

**Table 1.3.** Composite International Diagnostic Inventory (CIDI) Depression UK Biobank Questions and Field Codes

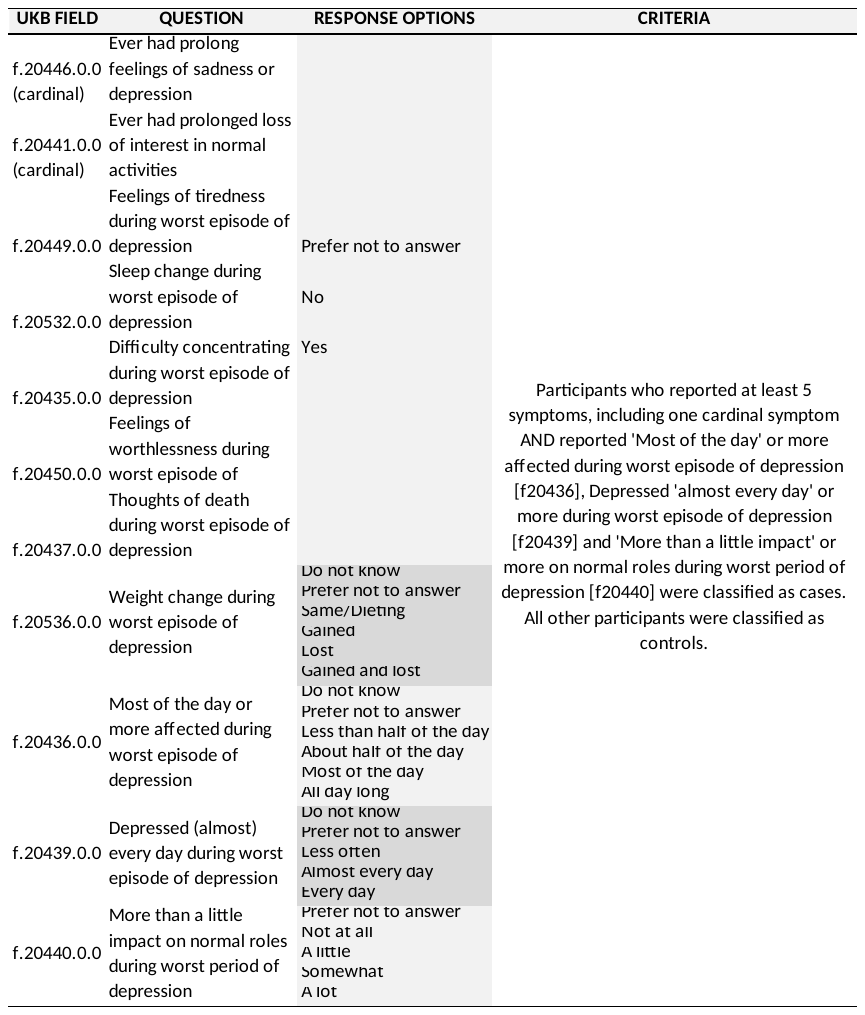

**Table 1.4.** Exclusion Criteria UK Biobank Questions and Field Codes

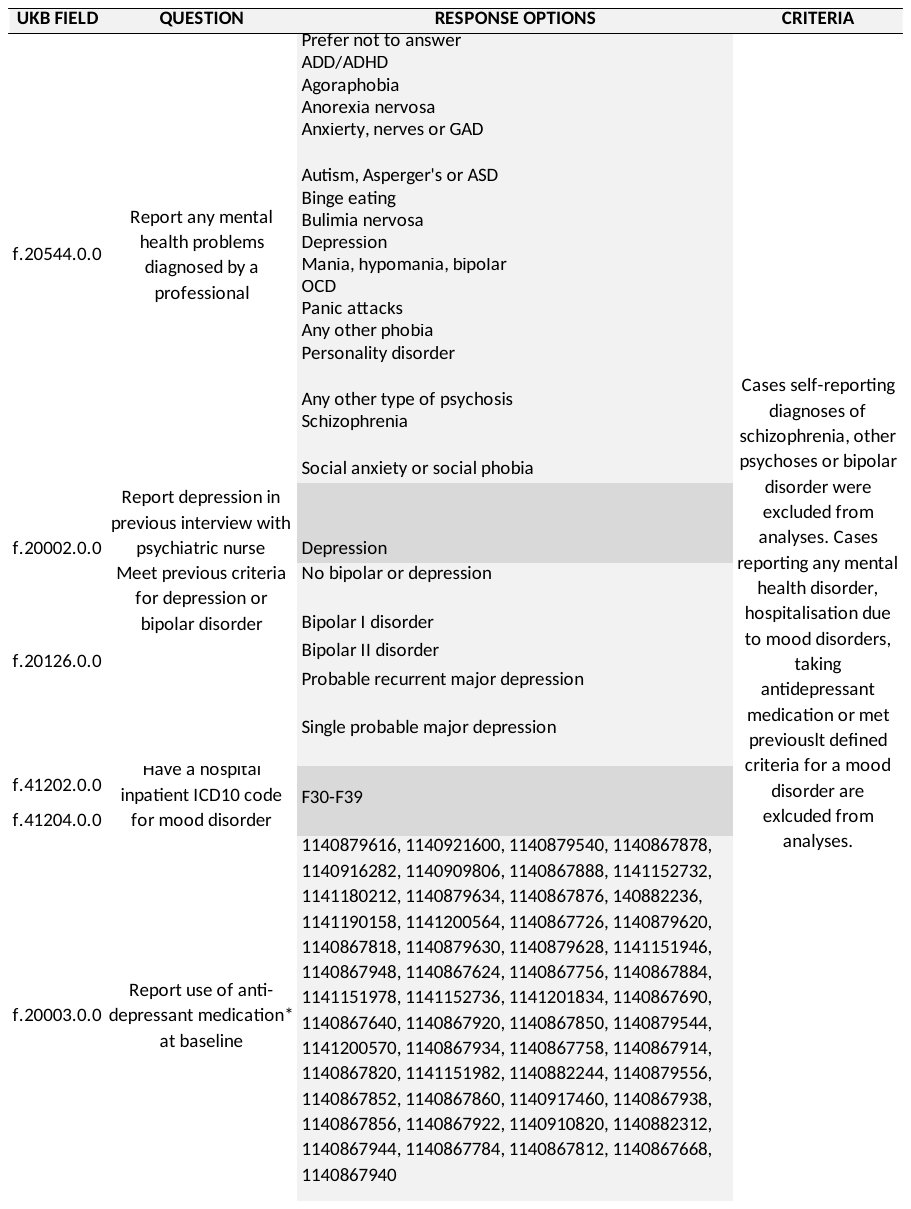

**Table 2** presents associations between UK Biobank trauma exposure questions and the phenotypes of interest; broad depression, CIDI depression and neuroticism. Trauma exposure question responses were ‘Rarely True’, ‘Sometimes True’, ‘Often True’, ‘Always True’. Trauma exposure was explored as a categorical variable, and covariates; age and sex, were included in analyses. Table 2 was presented in separate sections for each trauma exposure sub-category (childhood, adult and catastrophic trauma exposure) and each phenotype of interest. Tables include β coefficient estimates of associations, standard errors, T and P-Values as well as phenotypic variance accounted for by each model.

**Table 2.1.1.** Childhood Trauma Exposure and CIDI Depression Regression Model Associations

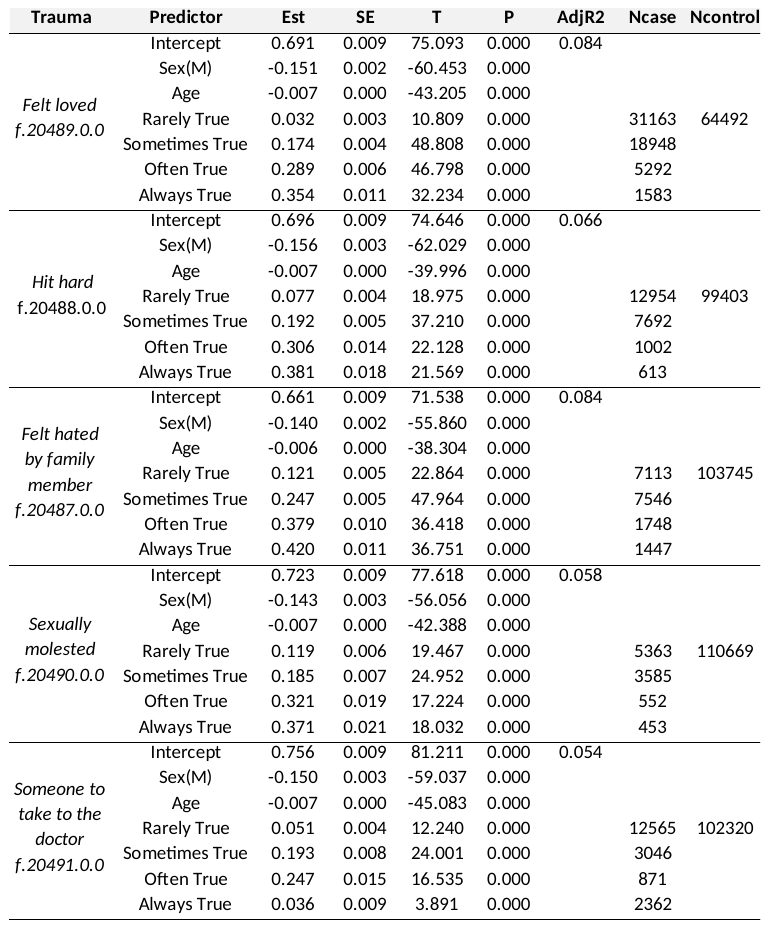

*Abbreviations.* CIDI, Composite International Diagnostic Inventory definition of depression; Sex(M), Male; Est, estimate; SE, standard error; T, T-Value; P, P-Value; AdjR2, adjusted R^2^ value; n, sample size.

**Table 2.1.2.** Childhood Trauma Exposure and Broad Depression Regression Model Associations

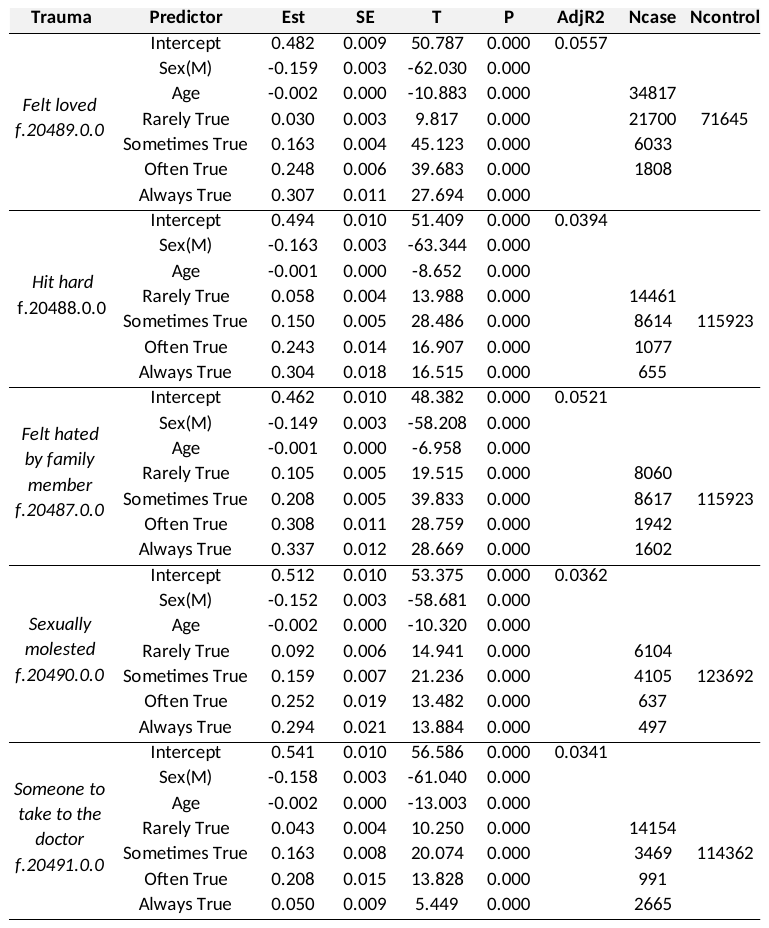

*Abbreviations.* Sex(M), Male; Est, estimate; SE, standard error; T, T-Value; P, P-Value; AdjR2, adjusted R^2^ value; n, sample size.

**Table 2.1.3.** Childhood Trauma Exposure and Neuroticism Regression Model Associations

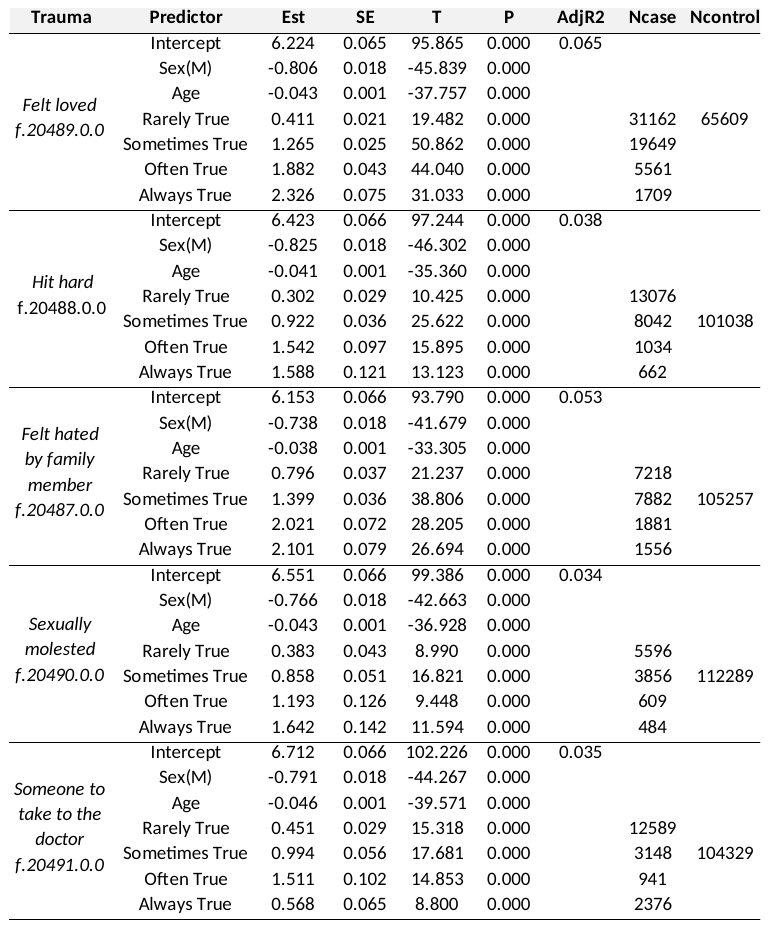

*Abbreviations.* Sex(M), Male; Est, estimate; SE, standard error; T, T-Value; P, P-Value; AdjR2, adjusted R^2^ value; n, sample size.

**Table 2.2.1.** Adult Trauma Exposure and CIDI Depression Regression Model Associations

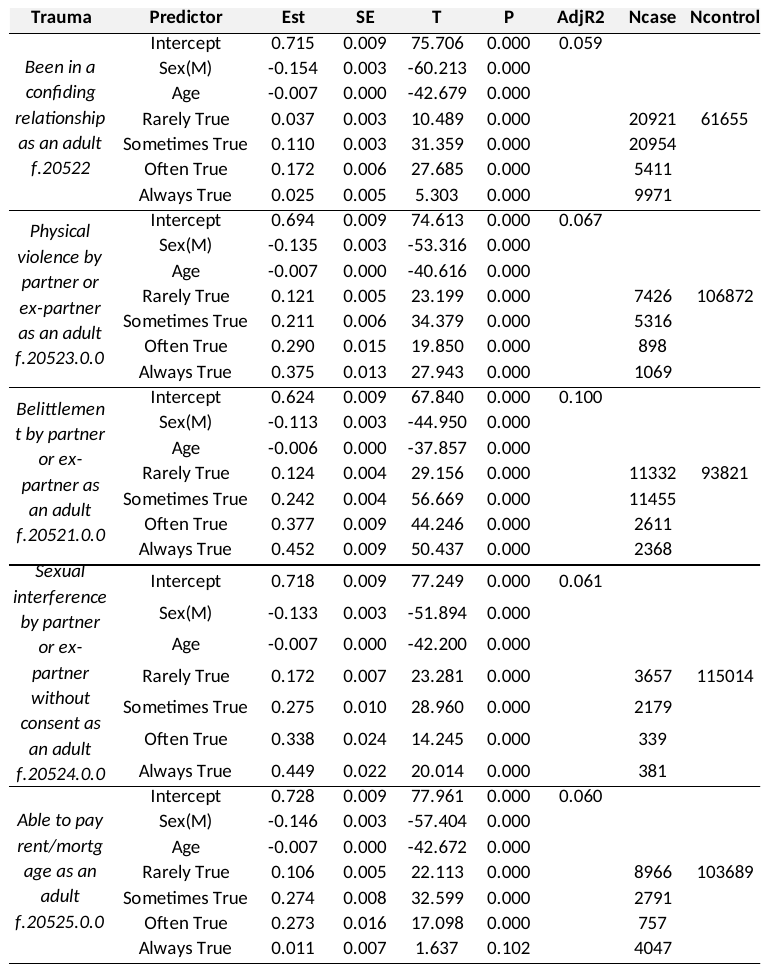

*Abbreviations.* CIDI, Composite International Diagnostic Inventory definition of depression; Sex(M), Male; Est, estimate; SE, standard error; T, T-Value; P, P-Value; AdjR2, adjusted R^2^ value; n, sample size.

**Table 2.2.2.** Adult Trauma Exposure and Broad Depression Regression Model Associations

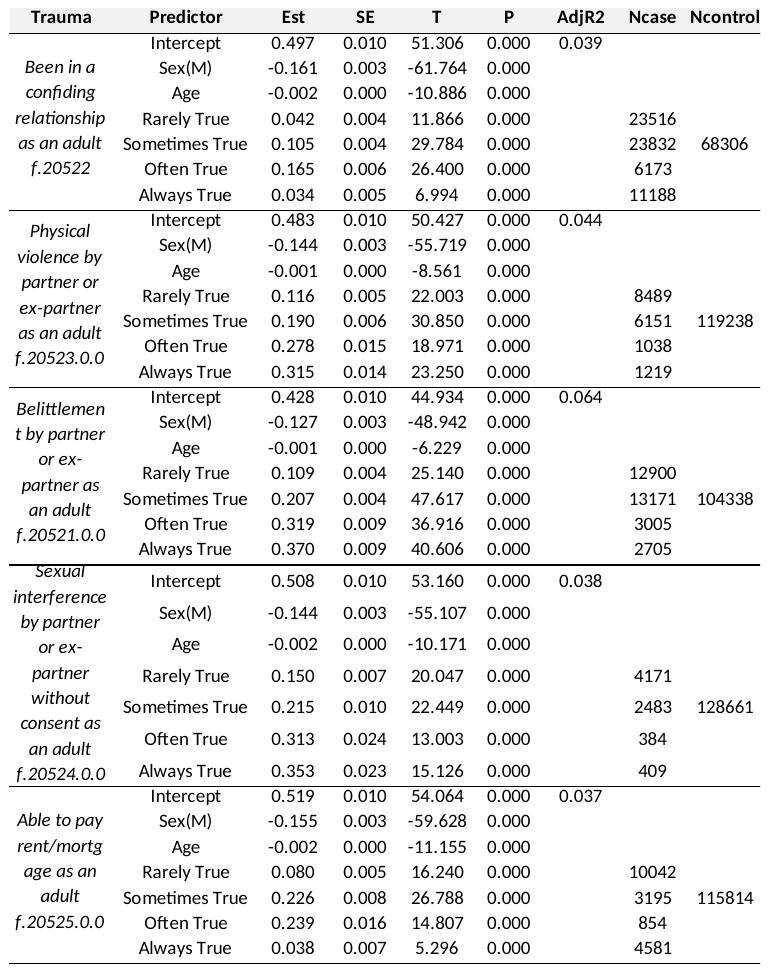

*Abbreviations.* Sex(M), Male; Est, estimate; SE, standard error; T, T-Value; P, P-Value; AdjR2, adjusted R^2^ value; n, sample size.

**Table 2.2.3.** Adult Trauma Exposure and Neuroticism Regression Model Associations

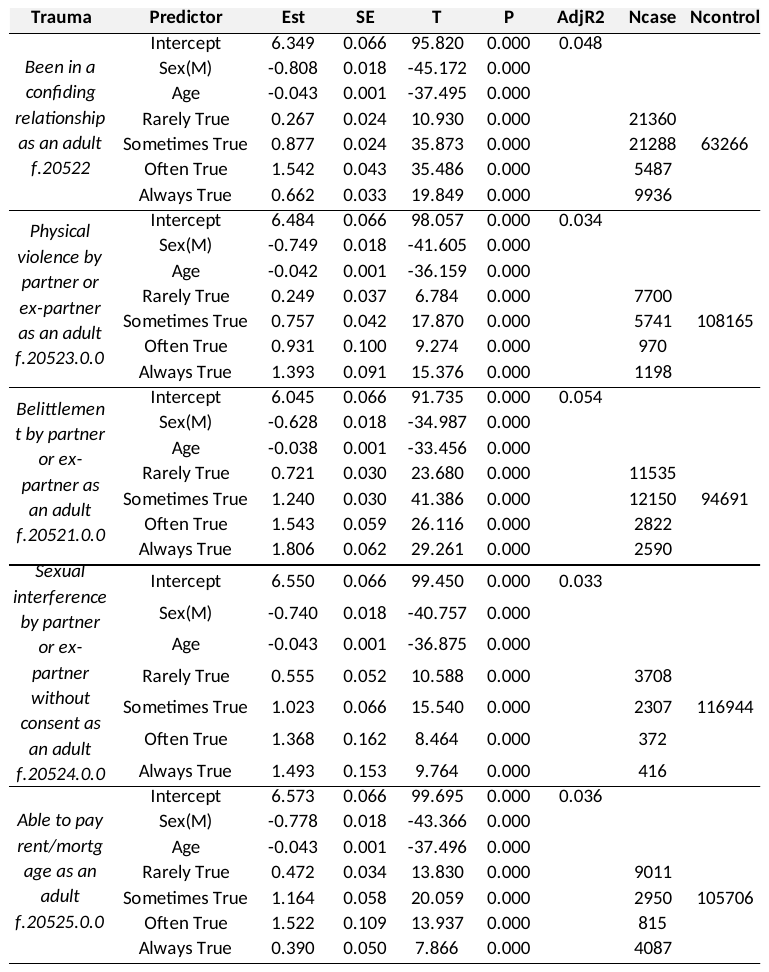

*Abbreviations.* Sex(M), Male; Est, estimate; SE, standard error; T, T-Value; P, P-Value; AdjR2, adjusted R^2^ value; n, sample size.

**Table 2.3.1.** Catastrophic Trauma Exposure and CIDI Depression Regression Model Associations

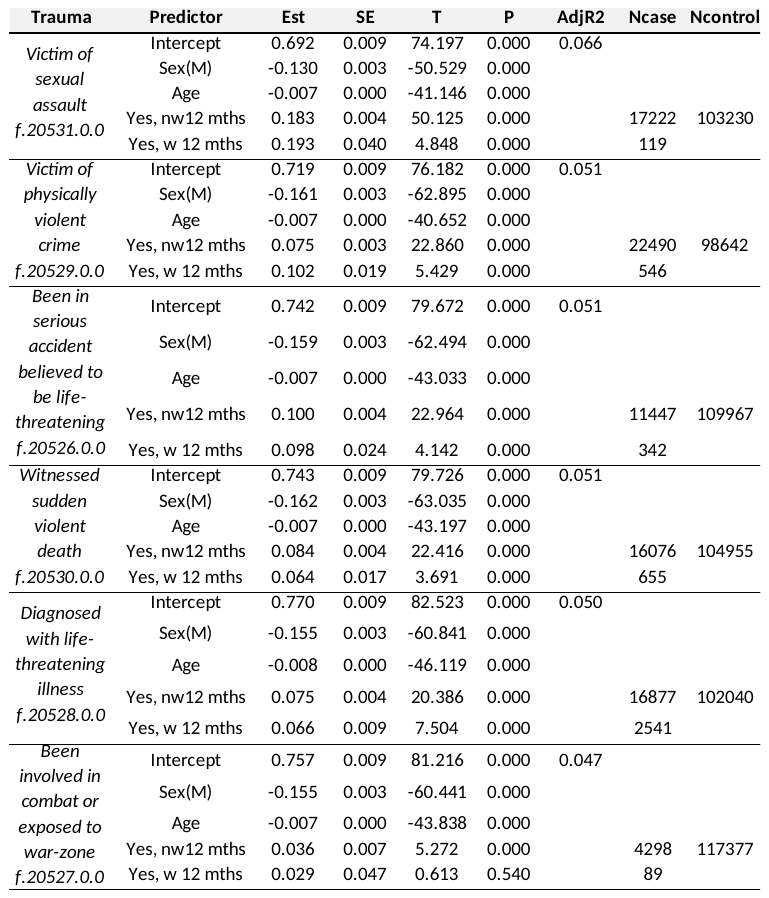

*Abbreviations.* CIDI, Composite International Diagnostic Inventory definition of depression; Sex(M), Male; Est, estimate; SE, standard error; T, T-Value; P, P-Value; AdjR2, adjusted R^2^ value; n, sample size.

**Table 2.3.2.** Catastrophic Trauma Exposure and Broad Depression Regression Model Associations

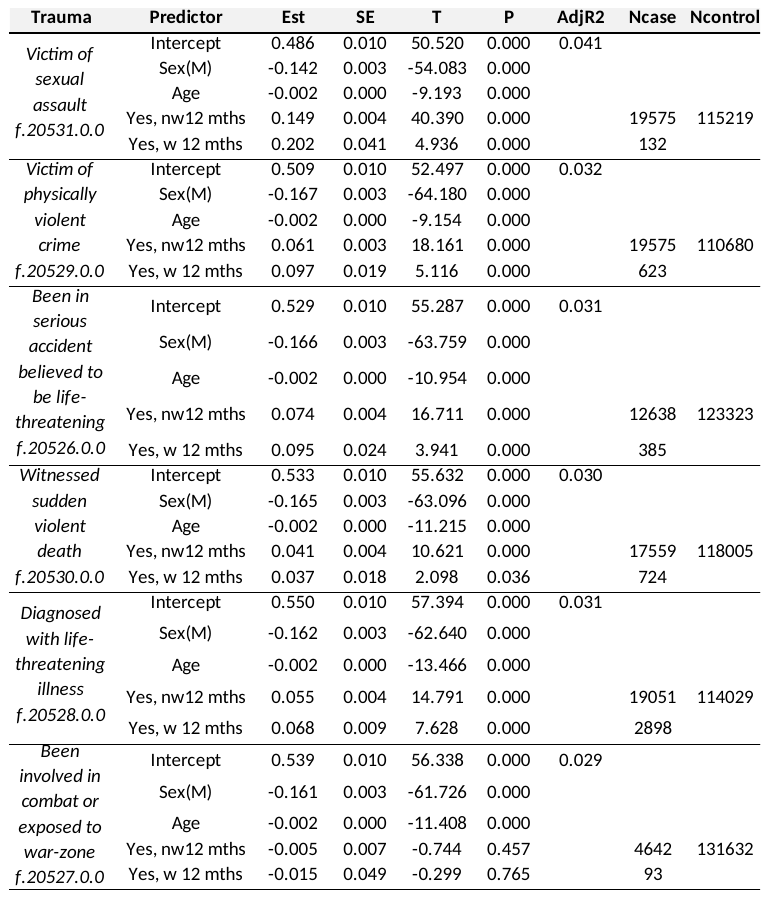

*Abbreviations.* Sex(M), Male; Est, estimate; SE, standard error; T, T-Value; P, P-Value; AdjR2, adjusted R^2^ value; n, sample size.

**Table 2.3.3.** Catastrophic Trauma Exposure and Neuroticism Regression Model Associations

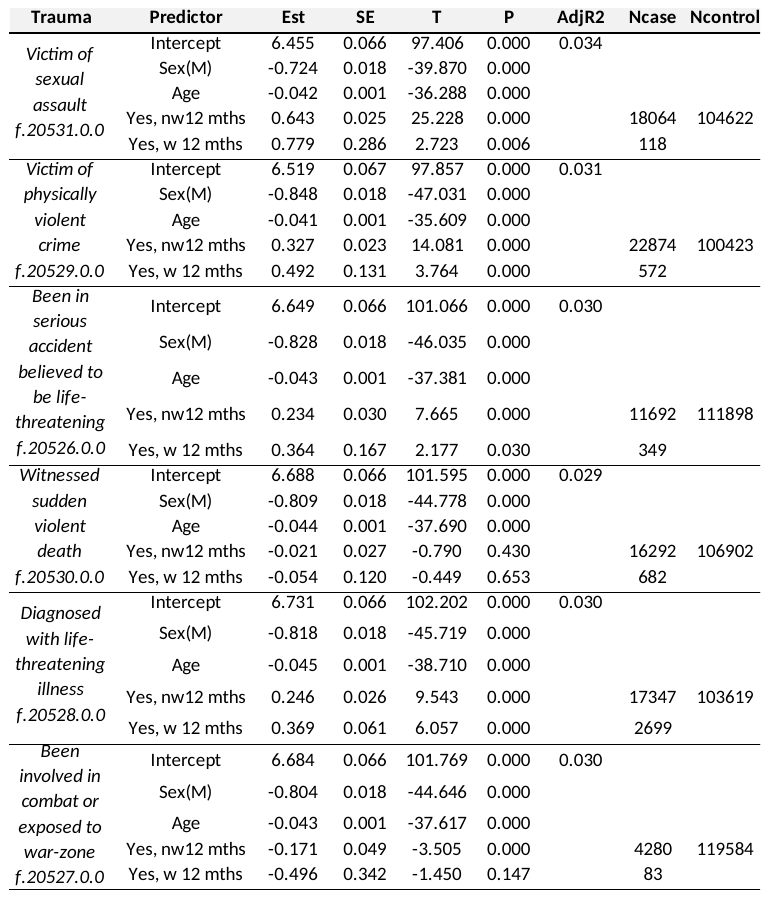

*Abbreviations.* Sex(M), Male; w/nw, within/not within; Est, estimate; SE, standard error; T, T-Value; P, P-Value; AdjR2, adjusted R^2^ value; n, sample size.

**Table 3** presents associations between UK Biobank trauma exposure eigenvectors, or also known as principal components (PCs) and the phenotypes of interest; broad depression, CIDI depression and neuroticism. Trauma PCs were obtained for full, childhood, adult and catastrophic trauma separately. Full trauma PCs pre-corrected for the full sample GRMs were also explored. Table 3 was presented in separate sections for each trauma exposure sub-category; full, childhood, adult and catastrophic trauma exposure. Tables include β coefficient estimates of associations, standard errors, T and P-Values as well as phenotypic variance accounted for by each model.

**Table 3.1.** Childhood Trauma Exposure Principal Components and Depression/Neuroticism Regression Model Associations

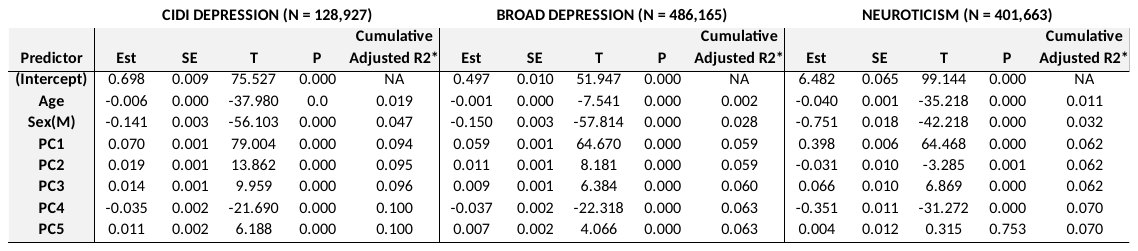

*Abbreviations.* CIDI, Composite International Diagnostic Inventory definition of depression; PC, principal component; Sex(M), Male; Est, estimate; SE, standard error; T, T-Value; P, P-Value; AdjR2; N, sample size. Coefficients are obtained from the full multiple regression model (including all predictors). *Represent adjusted R2 values when the specific predictor is ADDED to the model, on top of prior predictors.

**Table 3.2.** Adult Trauma Exposure Principal Components and Depression/Neuroticism Regression Model Associations

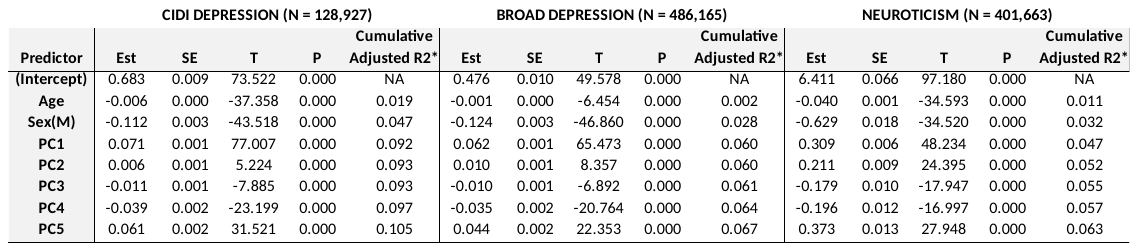

*Abbreviations.* CIDI, Composite International Diagnostic Inventory definition of depression; PC, principal component; Sex(M), Male; Est, estimate; SE, standard error; T, T-Value; P, P-Value; N, sample size. Coefficients are obtained from the full multiple regression model (including all predictors). *Represent adjusted R2 values when the specific predictor is ADDED to the model, on top of prior predictors.

**Table 3.3.** Catastrophic Trauma Exposure Principal Components and Depression/Neuroticism Regression Model Associations

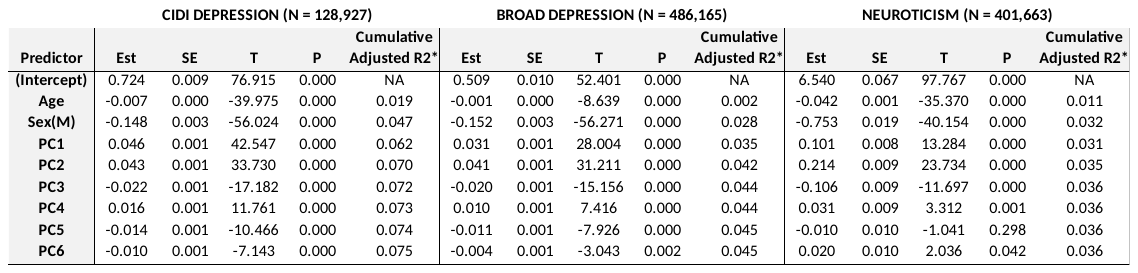

*Abbreviations.* CIDI, Composite International Diagnostic Inventory definition of depression; PC, principal component; Sex(M), Male; Est, estimate; SE, standard error; T, T-Value; P, P-Value; N, sample size. Coefficients are obtained from the full multiple regression model (including all predictors). *Represent adjusted R2 values when the specific predictor is ADDED to the model, on top of prior predictors.

**Table 3.4.** Full Trauma Exposure Principal Components and Depression/Neuroticism Regression Model Associations

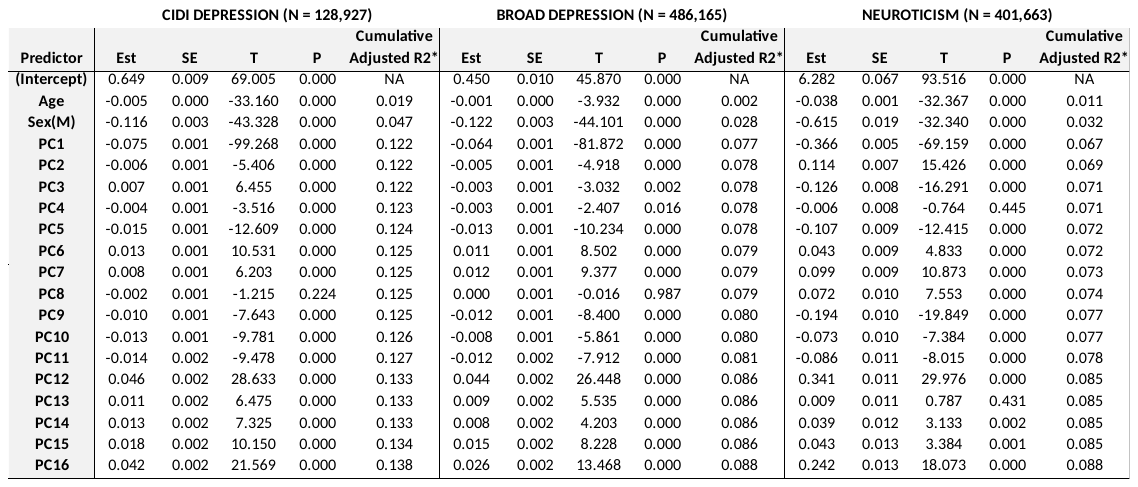

*Abbreviations.* CIDI, Composite International Diagnostic Inventory definition of depression; PC, principal component; Sex(M), Male; Est, estimate; SE, standard error; T, T-Value; P, P-Value; N, sample size. Coefficients are obtained from the full multiple regression model (including all predictors). *Represent adjusted R2 values when the specific predictor is ADDED to the model, on top of prior predictors.

**Table 4** presents trauma exposure principal component (PC) loadings. Here, values represent how each trauma exposure question loads on to the separate PCs. Loadings are presented separately for each trauma exposure sub-category; full, childhood, adult and catastrophic trauma exposure. Table 4 is presented in separate sections for each trauma exposure sub-category.

**Table 4.1.** Childhood Trauma Exposure Principal Component Loadings

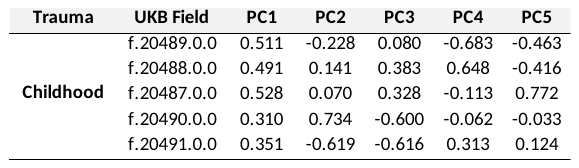

*Abbreviations.* UKB, UK Biobank; PC, principal component.

**Table 4.2.** Adult Trauma Exposure Principal Component Loadings

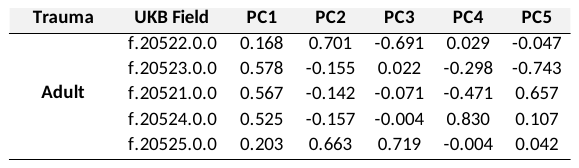

*Abbreviations.* UKB, UK Biobank; PC, principal component.

**Table 4.3.** Catastrophic Trauma Exposure Principal Component Loadings

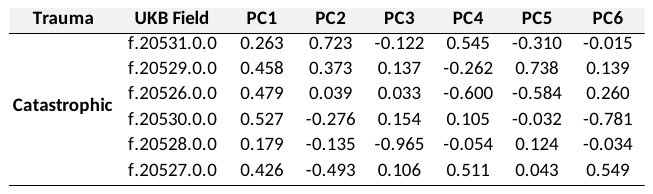

*Abbreviations.* UKB, UK Biobank; PC, principal component.

**Table 4.4.** Full Trauma Exposure Principal Component Loadings

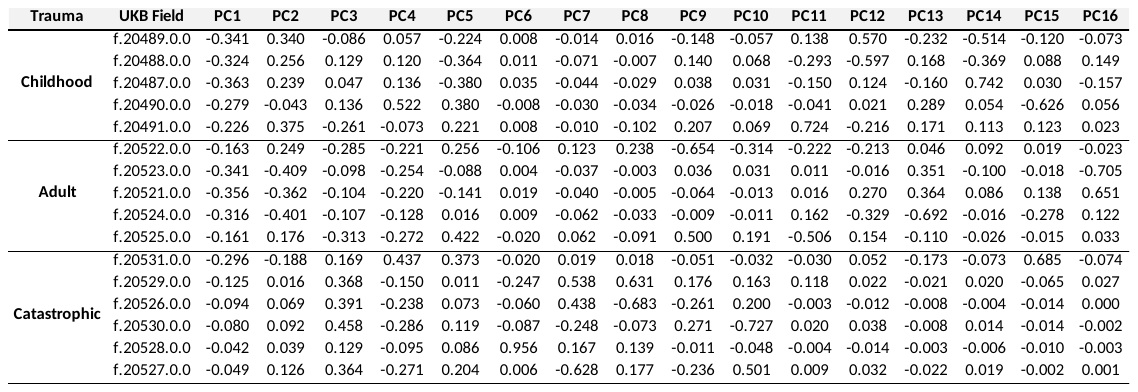

*Abbreviations.* UKB, UK Biobank; PC, principal component.

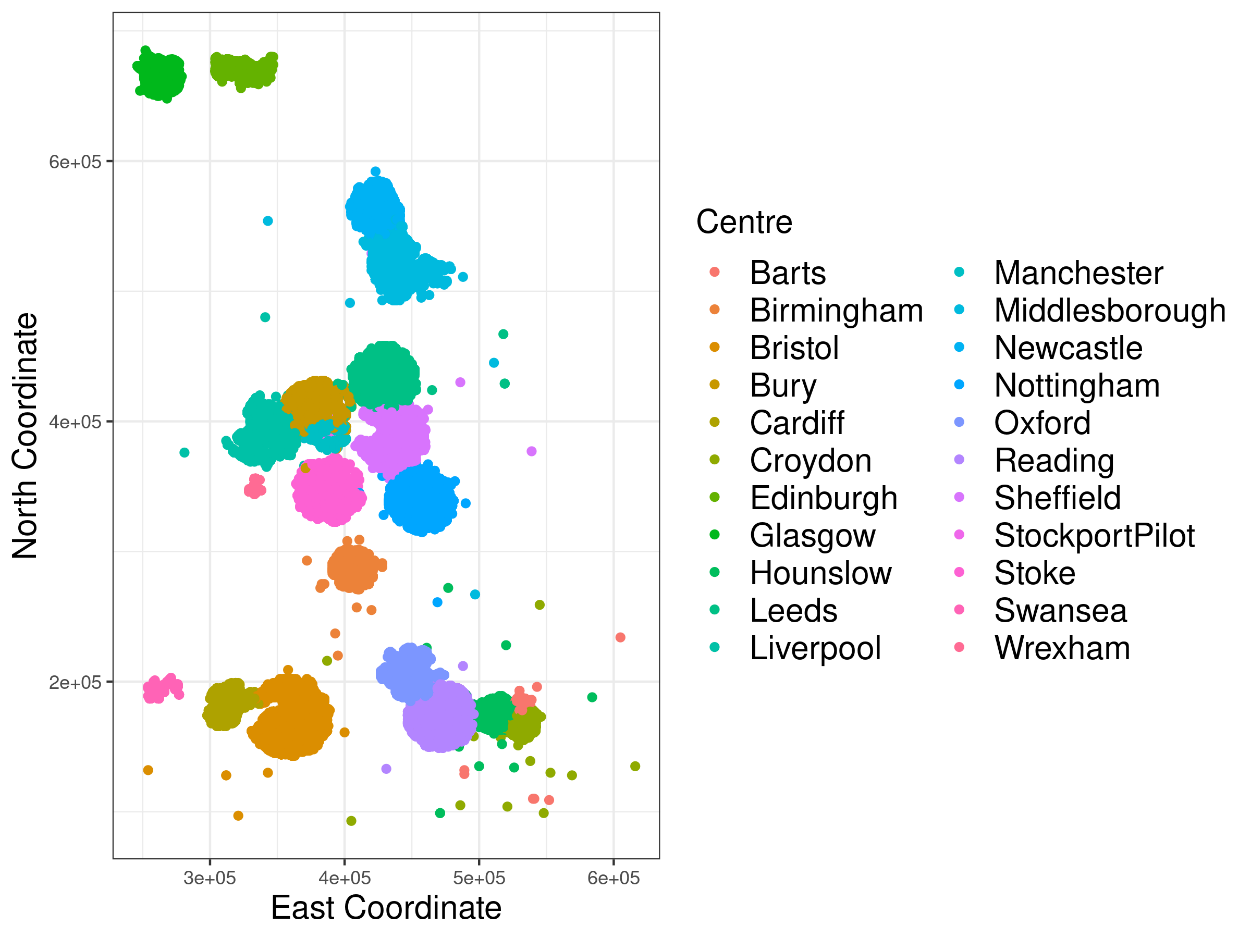

| **Clusters** | **Centre** | **Sample Size** |
| --- | --- | --- |
| **North** | Glasgow | 4503 |
|  | Edinburgh | 5592 |
|  | Newcastle | 9771 |
|  | Middlesborough | 6214 |
| **Mid North** | Leeds | 12332 |
|  | Bury | 6915 |
|  | Manchester | 3982 |
|  | Liverpool | 8969 |
| **Mid South** | Wrexham | 151 |
|  | Stoke | 4304 |
|  | StockportPilot | 294 |
|  | Sheffield | 9290 |
|  | Nottingham | 10193 |
|  | Birmingham | 6945 |
| **South West** | Swansea | 660 |
|  | Cardiff | 4731 |
|  | Bristol | 14642 |
| **South East** | Oxford | 4897 |
|  | Reading | 10904 |
|  | Hounslow | 9542 |
|  | Croydon | 8860 |
|  | Barts | 4438 |

***Figure 2*.** UK Biobank Geographical Clusters. Above, figure shows the North and East co-ordinates of participant residence and colours represent the recruitment centres for each participant. Below, table shows geographical clusters, respective centres and sample sizes.

**Table 5** presents demographic information on individuals within each geographical cluster; North, Mid-North, Mid-South, South-West, South-East. Demographic information for each cluster was presented when clusters were formed using all individuals responding to ALL trauma exposure questions. Additional demographic information tables were provided for only unrelated individuals responding to all trauma exposure questions as well as all individuals who have responded to all childhood, adult and catastrophic trauma exposure questions separately. Table 5 was presented in separate sections for each sample.

**Table 5.1.** Cluster Demographics Using All Participants With Complete Trauma Exposure Responses

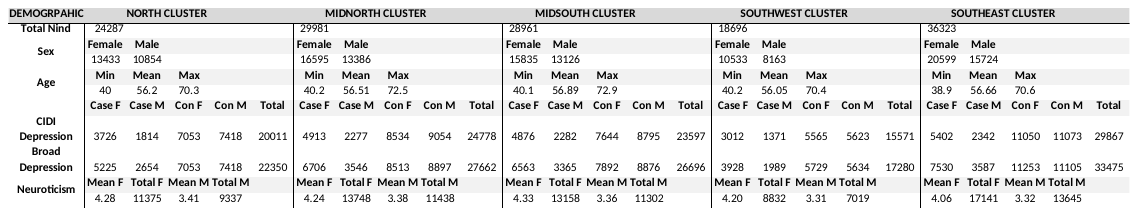

*Abbreviations.* Nind, number of individuals; Min, minimum; Max, maximum; F, M, Female and Male, respectively.

**Table 5.2.** Cluster Demographics Using Unrelated Participants With Complete Trauma Exposure Responses

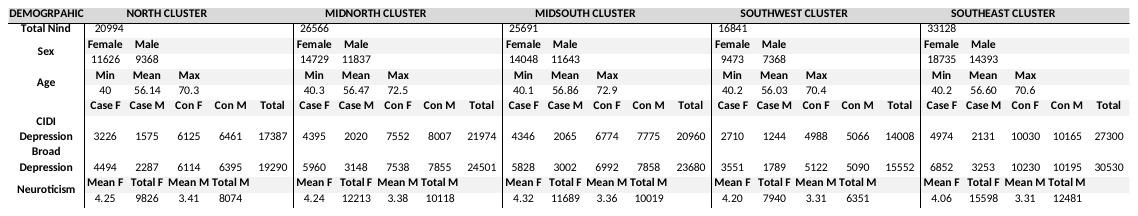

*Abbreviations.* Nind, number of individuals; Min, minimum; Max, maximum; F, M, Female and Male, respectively.

**Table 5.3.** Cluster Demographics Using All Participants With Complete Childhood Trauma Exposure Responses

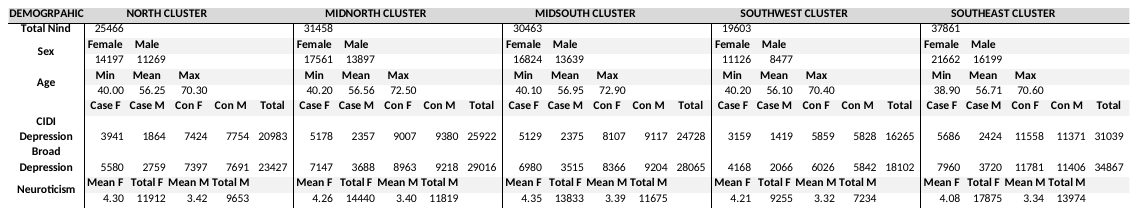

*Abbreviations.* Nind, number of individuals; Min, minimum; Max, maximum; F, M, Female and Male, respectively.

**Table 5.4.** Cluster Demographics Using All Participants With Complete Adult Trauma Exposure Responses

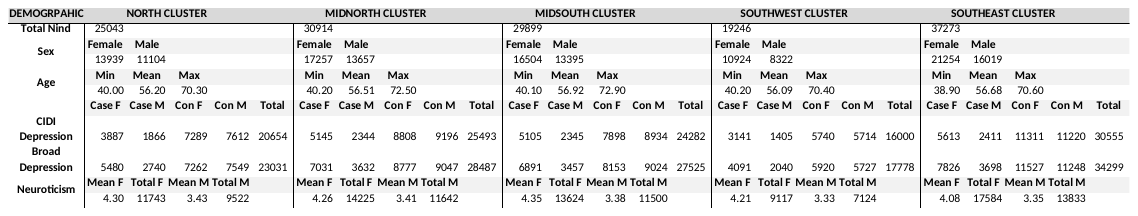

*Abbreviations.* Nind, number of individuals; Min, minimum; Max, maximum; F, M, Female and Male, respectively.

**Table 5.5.** Cluster Demographics Using All Participants With Catastrophic Trauma Exposure Responses

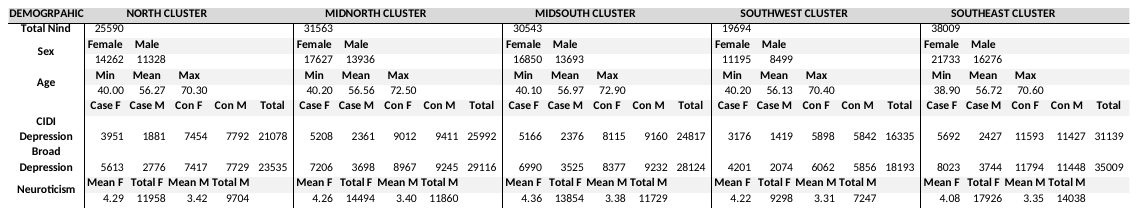

*Abbreviations.* Nind, number of individuals; Min, minimum; Max, maximum; F, M, Female and Male, respectively.

**Table 6** presents demographic information on individuals within each geographical cluster; North, Mid-North, Mid-South, South-West, South-East. Demographic information for each cluster was presented when clusters were formed using all individuals responding to ALL trauma exposure questions. Table 6 was presented in separate sections for each sex.

**Table 6.1.** Cluster Demographics Using All Female Participants With Complete Trauma Exposure Responses

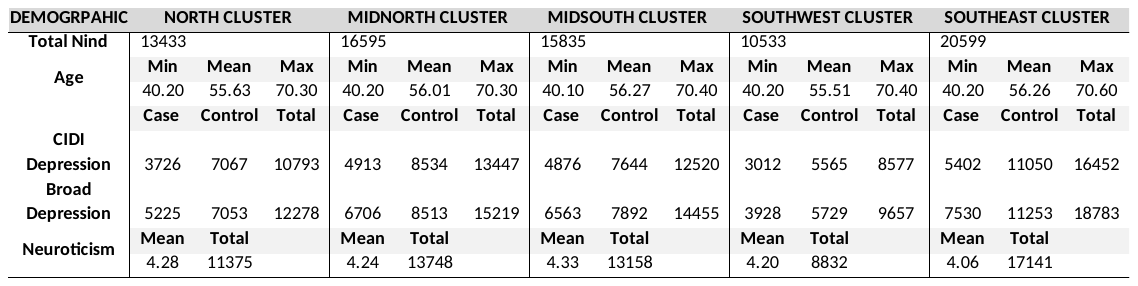

*Abbreviations.* Nind, number of individuals; Min, minimum; Max, maximum.

**Table 6.2.** Cluster Demographics Using Unrelated Female Participants With Complete Trauma Exposure Responses

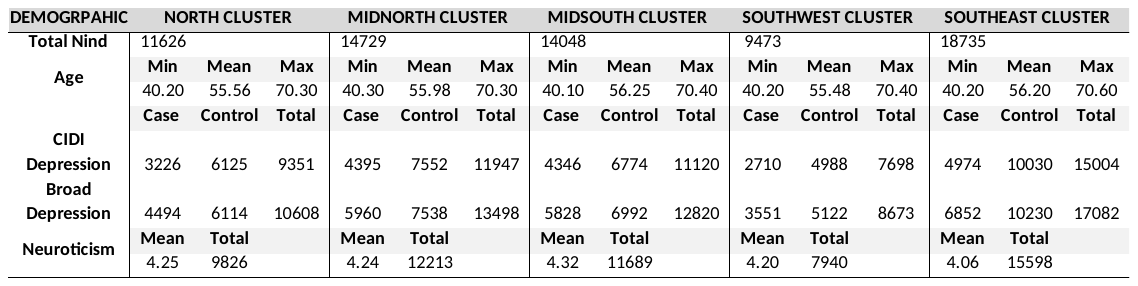

*Abbreviations.* Nind, number of individuals; Min, minimum; Max, maximum.

**Table 6.3.** Cluster Demographics Using All Male Participants With Complete Trauma Exposure Responses

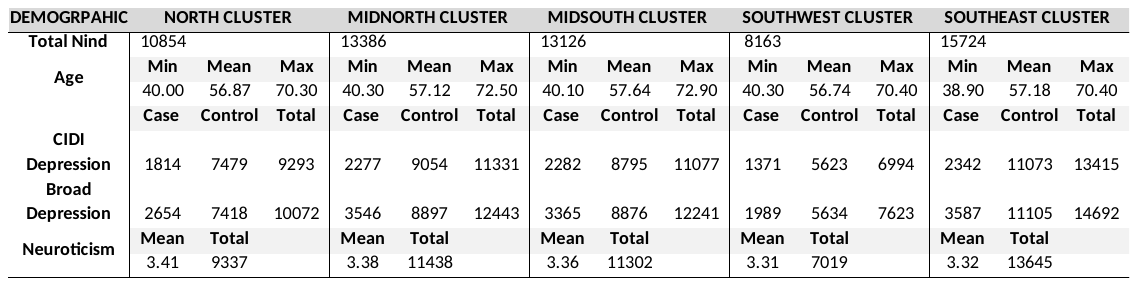

*Abbreviations.* Nind, number of individuals; Min, minimum; Max, maximum.

**Table 6.4.** Cluster Demographics Using Unrelated Male Participants With Complete Trauma Exposure Responses

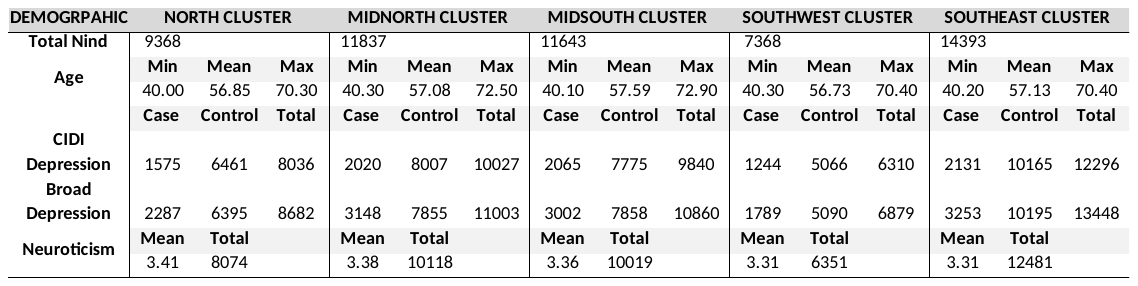

*Abbreviations.* Nind, number of individuals; Min, minimum; Max, maximum.

**Section A**

OmicS-data-based Complex-trait Analysis [OSCA v0.45](https://yanglab.westlake.edu.cn/software/osca/#Overview) is a software used to analyse available omics (e.g. DNA methylation) data (Zhang et al, 2019). Three separate equations are provided to compute Omics Relationship Matrices (ORMs). In this study, we refer to ORMs as environmental relationship matrices (Es), as this is more appropriate for the data being used in this study (trauma principal components).

Algorithm 1 computes similarity by using standardised DNAm measures of all probes. Here it is important to note that the DNAm measures will be replaced with the principal component eigenvectors (PCs) of our trauma exposure measures.

$$\mathbf{Equation 1}. A_{jk}= \Sigma_{i}\left( x_{ij}- \mu_{i} \right)\left( x_{ik}- \mu_{i} \right) / \sigma_{i}^{2}$$

With $A_{jk}$ being the environmental similarity between individuals $j$ and $k$. Where $x_{ij}$ and $x_{ik}$ is the **standardised** $\mathrm{PC}_{i}$ in individual $j$ and $k$, respectively. $\mu_{i}$ and $\sigma_{i}^{2}$ are the mean and variance of $\mathrm{PC}_{i}$ over all the individuals respectively.

Algorithm 2 computes similarity by using **unstandardised** eigenvectorss and factors in the number of eigenvectors utilised. This algorithm implicitly assumes eigenvectors with smaller variance tend to have larger effects on the phenotype, and that there is no relationship between the depression/neuroticism phenotypic variance captured by the eigenvector and the variance of the eigenvector. Here, we cannot confirm these assumptions, so equation 2, was deemed inappropriate for the data we have.

$$\mathbf{Equation 2}. A_{jk}= \frac{1}{m}\Sigma_{i}\left( x_{ij}- \mu_{i} \right)\left( x_{ik}- \mu_{i} \right) / \sigma_{i}^{2}$$

With $A_{jk}$ being the environmental similarity between individuals $j$ and $k$. Where $x_{ij}$ and $x_{ik}$ is the **unstandardised** $\mathrm{PC}_{i}$ in individual $j$ and $k$, respectively. $\mu_{i}$ and $\sigma_{i}^{2}$ are the mean and variance of $\mathrm{PC}_{i}$ over all the individuals respectively and $m$ is the number of PCs utilised.

Algorithm 3 computes similarity by iteratively standardising eigenvectors and across individuals.

$$\mathbf{Equation 3}. A_{jk}= \frac{1}{m}\Sigma_{i}\left( x_{ij}- \mu_{i} \right)\left( x_{ik}- \mu_{i} \right) / \Sigma_{i}\sigma_{i}^{2}$$

With $A_{jk}$ being the environmental similarity between individuals $j$ and $k$. Where $x_{ij}$ and $x_{ik}$ is the **standardised** $\mathrm{PC}_{i}$ in individual $j$ and $k$, respectively. $\mu_{i}$ and $\sigma_{i}^{2}$ are the mean and variance of $\mathrm{PC}_{i}$ over all the individuals respectively.

For our study, all algorithms were separately used to compute Es from full trauma eigenvectors (PCs), algorithm 1 and 3 were deemed the most appropriate for our data. Subtle differences were observed in outputted relationship matrices (**Figure 1**). Matrices computed using algorithm 1 present diagonal values with a mean of 1, whereas matrices computed using algorithm 3 present diagonal values of 1.

| 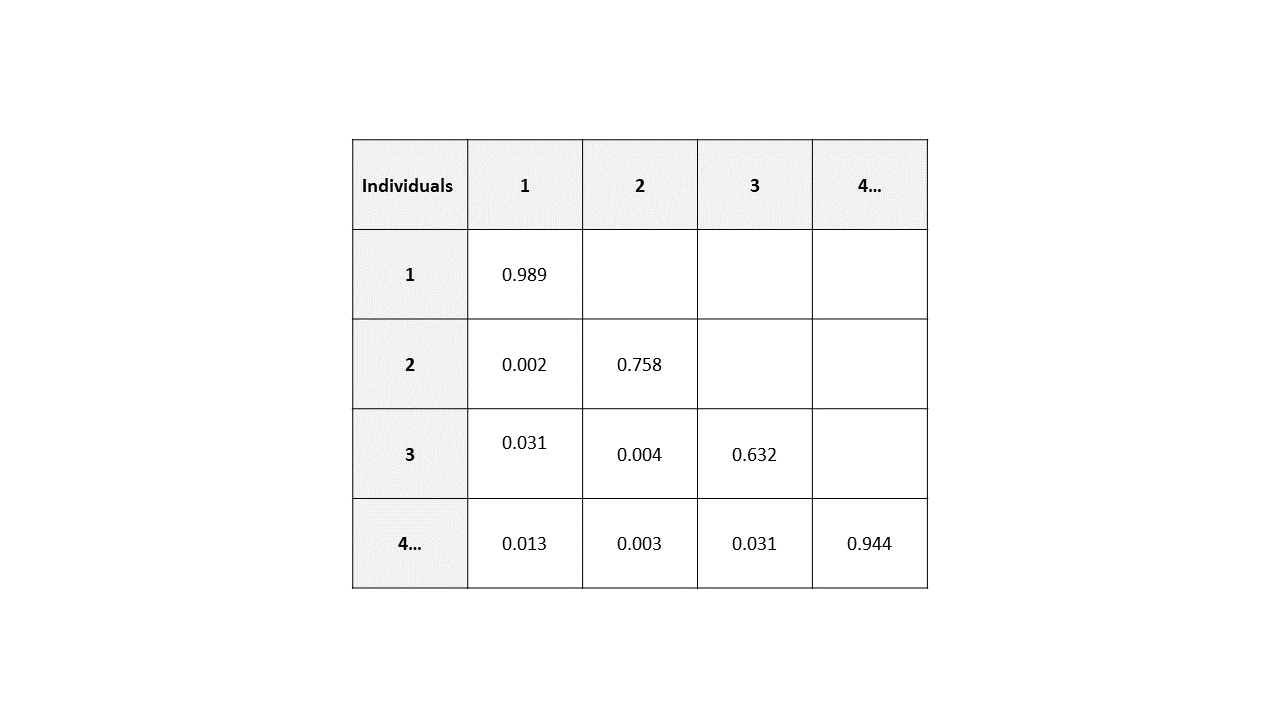 | 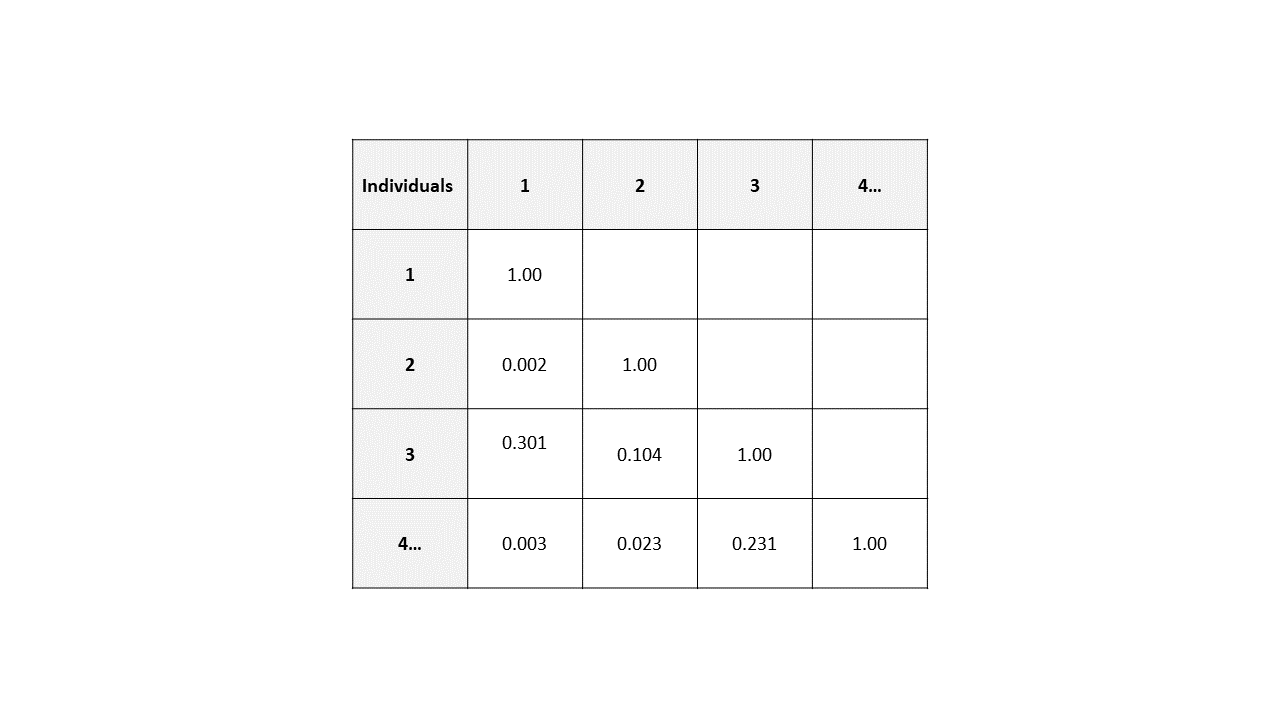 |
| --- | --- |
| ***Figure SA1.*** Trauma Exposure Relationship Matrices. Matrix diagonals represent similarity between an individual with themselves. Offdiagonals represent pairwise similarity between individuals within the sample. Here, similarity is computed using available trauma exposure principal components using OSCA software. Left, shows a representative similarity matrix where similarity is computed using Algorithm 1. Right, shows a representative similarity matrix where similarity is computed using Algorithm 3. Please note values specified are not obtained from real matrices. | |

Offdiagonal values of the computed Es and genomic relationship matrices (Gs) were plotted to visualise the relationship between values (**Figure 2**).

| 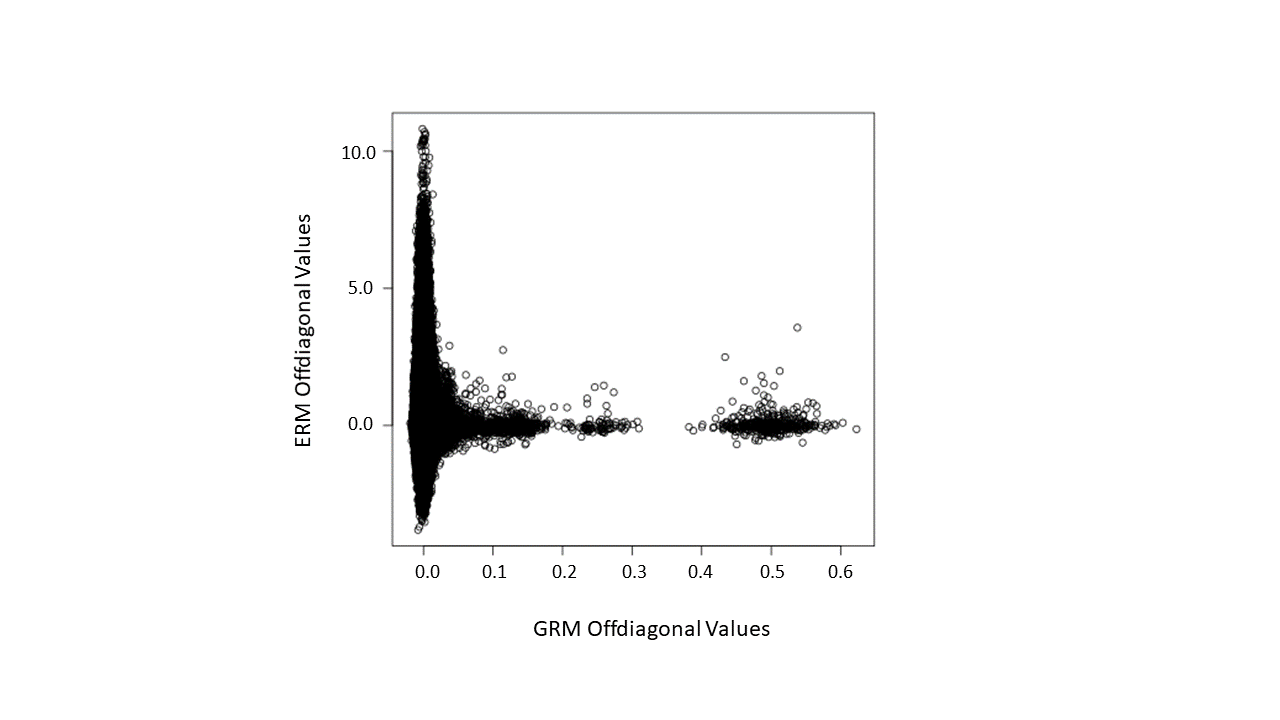 | 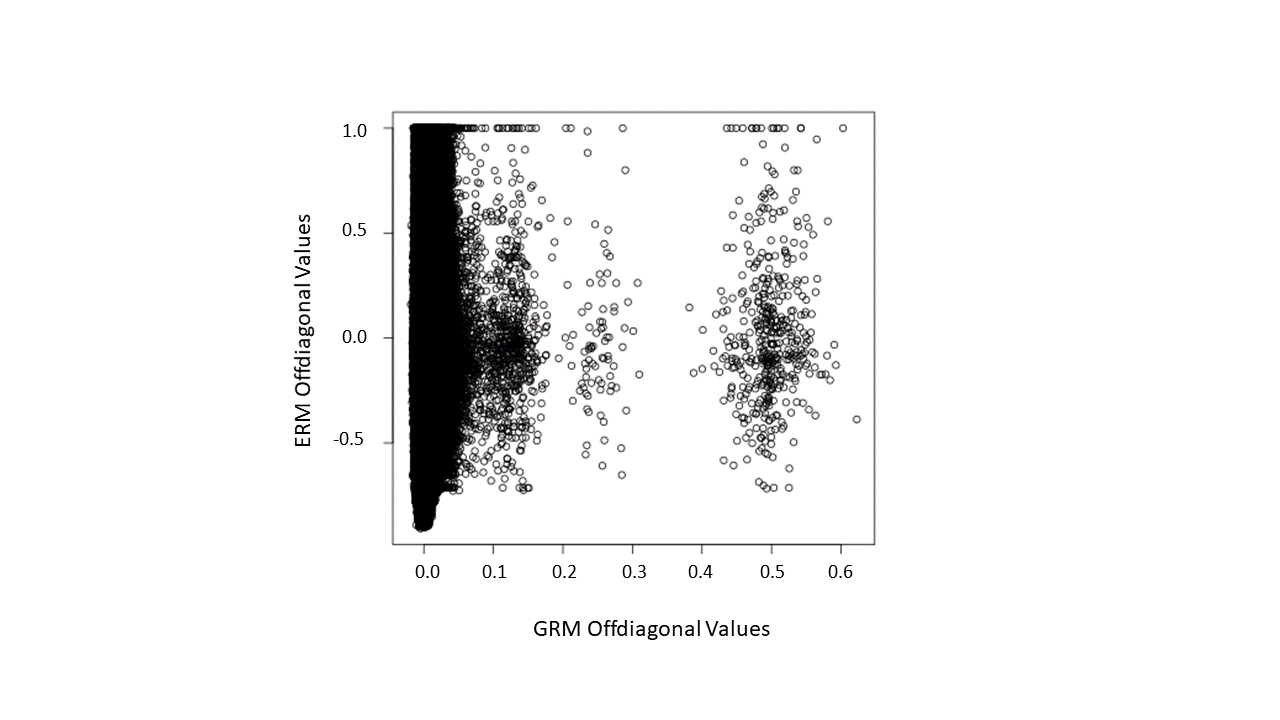 |
| --- | --- |
| ***Figure SA2*.** Offdiagonal Values of UKB North Geographical Cluster Genomic and Environmental Relationship Matrices. Left, Environmental Relationship Matrix (ERM) computed using Algorithm 1 of OSCA software. Right, ERM computed using Algorithm 3 of OSCA software. The x-axis represent offdiagonal genomic similarity values, cluster points surrounding points 0.2-0.3 and 0.5 show points of relatives. The y-axis represents offdiagonal trauma exposure similarity values. | |

Zhang and colleagues (2019) demonstrated, with simulation analyses, that mixed linear model anlaysis results obtained from matrices computed with algorithm 1 and 3 showed minimal differences. Similarly, we show that GREML outputs were negligibly different between the algorithms of interest (**Table 7**).

Here, algorithm 1 is opted when computing the ERMs used for downstream analyses. The mixed linear model results are similar and the plots from **figure 2** suggest values in line with expectations.

**Table 7** presents results of mixed linear models utilising genomic relationship matrices, trauma exposure relationship matrices and genome-by-trauma exposure relationship matrices as random effects. Results include estimates of CIDI depression variance attributable to each random effect, standard errors, log-ratio test values and log-ratio test p-values signifying model utility. Table 7 presents results of mixed linear models whereby trauma exposure relationship matrices were computed using the 3 different algorithms available within OmicS-data-based Complex-trait Analysis OSCA software.

**Table 7.** Mixed Linear Model Results of Proportion of CIDI Depression Variance Attributable to Environmental Relationship Matrices Computed Using Different OSCA Algorithms

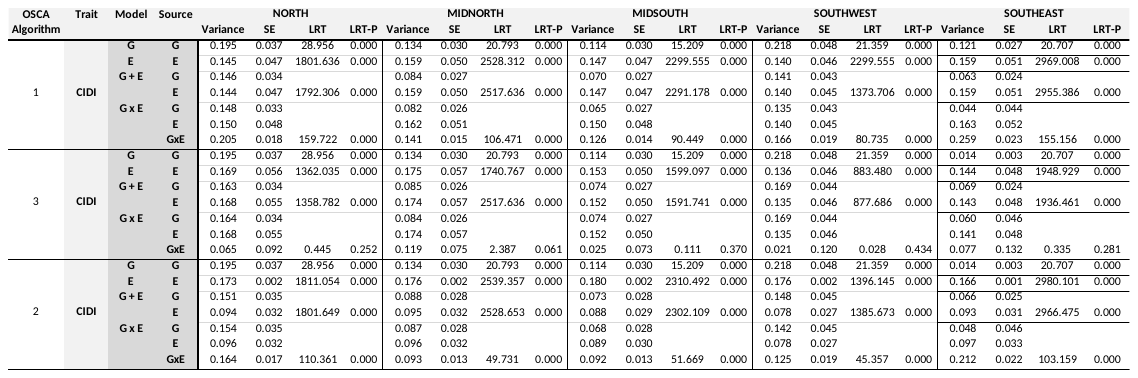

*Abbreviations.* CIDI, Composite International Diagnostic Inventory Depression; OSCA, OmicS-based-data Complex-trait Analysis; G, Genomic Relationship Matrix; E, Environmental (trauma exposure) Relationship Matrix; GxE, Genome-by-Trauma Exposure Interaction Relationship Matrix; SE, standard error; LRT, log-ratio test; P, P-Value.

**Table 8** presents heritability estimates obtained for trauma exposure obtained from Haseman-Elston regression models. The table presents estimates of variance attributable to the genomic relationship matrix, standard errors and P-Values from analyses conducted on the whole and unrelated samples separately. Heritability estimates were obtained separately for full trauma, childhood, adult and catastrophic trauma exposure.

**Table 8.** Trauma Exposure Heritability Estimates

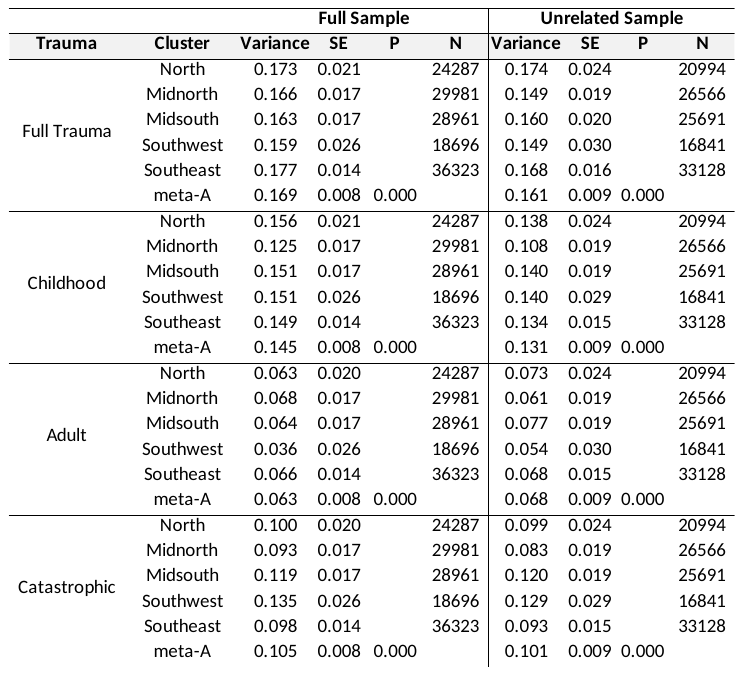

*Abbreviations.* SE, standard error, P, P-Value; N, sample size; meta-A, Meta-analysed estimates of variance and SEs.

**Table 9** presents estimates of genetic correlations between trauma exposure and depression/neuroticism variables. Tables present estimates, standard errors and P-Values.

**Table 9.** Trauma Exposure and Depression/Neuroticism Genetic Correlations

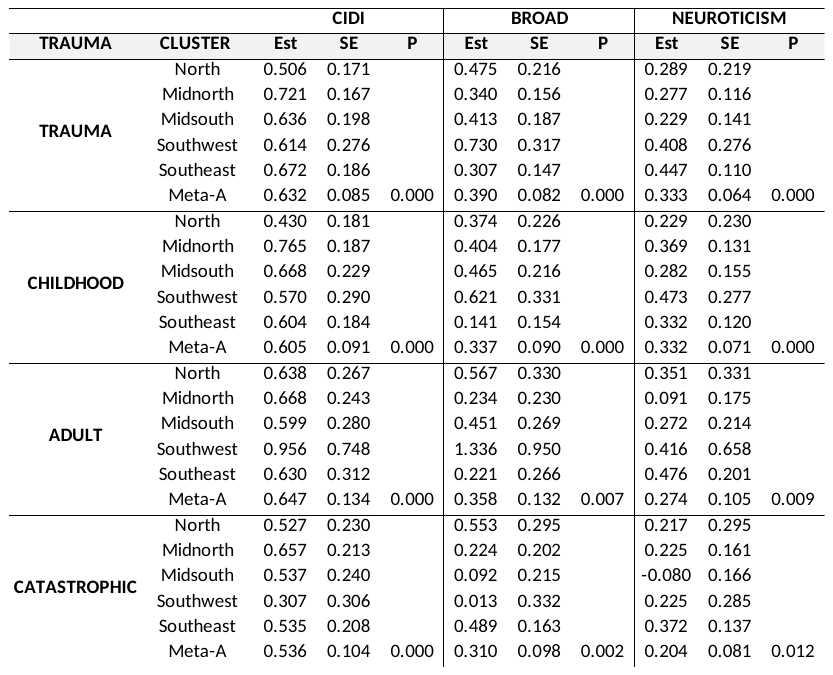

*Abbreviations.* CIDI, Composite International Diagnostic Inventory definition of depression; Est, estimate; SE, standard error; P, P-Value; Meta-A, meta analysed values of estimates and SEs.

**Tables 10-22** presents results of mixed linear models using genomic relationship matrices (Gs), environmental relationship matrices (Es) using trauma exposure eigenvectors (PCs), and genome-by-trauma exposure interaction relationship matrices (GxE) as random effects for each geographical cluster. Results include estimates of trait variance attributable to the G, E and GxE, standard errors and P-Values. Results also include meta-analysed estimates and standard errors. Each table presents results of models utilising either different samples (unrelated, female only, males only) or different Es (using full trauma exposure principal components PCs, childhood trauma exposure PCs, adult trauma exposure PCs etc.). Details of the samples and Es utilised are available in table headings.

**Table 10.** Mixed Linear Model Results Including Es of Full Trauma Exposure Principal Components.

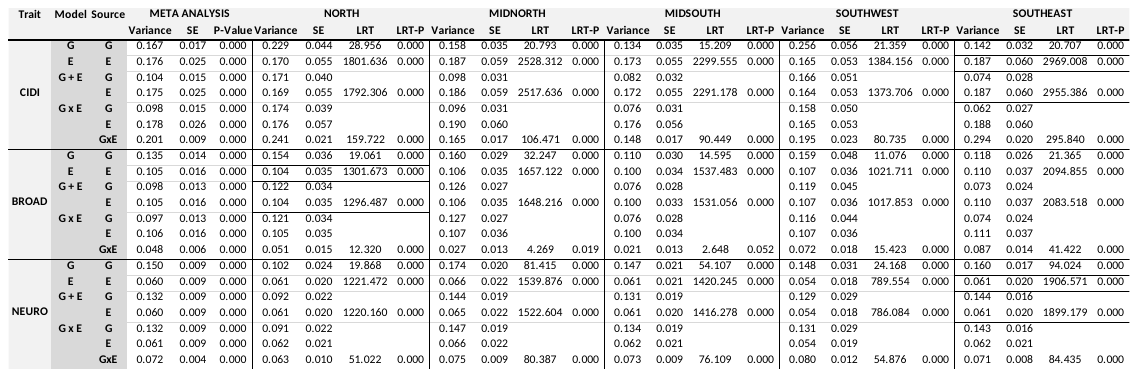

*Abbreviations.* E, environmental relationship matrix using full trauma principal components; SE, standard error; LRT, log-ratio test value; LRT-P, log-ratio test P-Value; G, genetic (Genomic Relationship Matrix); E, environmental (full trauma exposure ERM); GxE, genome-by-trauma exposure interaction.

**Table 11.** Mixed Linear Model Results Including Es of Childhood Trauma Exposure Principal Components.

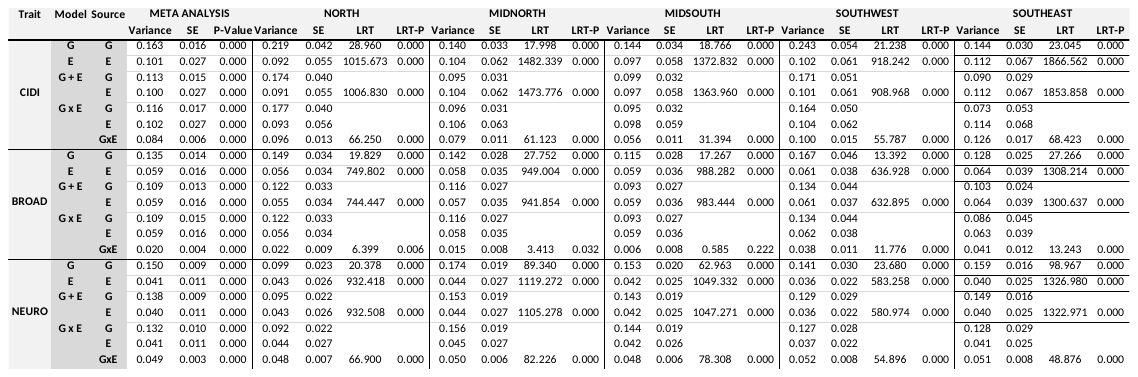

*Abbreviations.* E, environmental relationship matrix using full trauma principal components; SE, standard error; LRT, log-ratio test value; LRT-P, log-ratio test P-Value; G, genetic (Genomic Relationship Matrix); E, environmental (full trauma exposure ERM); GxE, genome-by-trauma exposure interaction.

**Table 12.** Mixed Linear Model Results Including Es of Adult Trauma Exposure Principal Components.

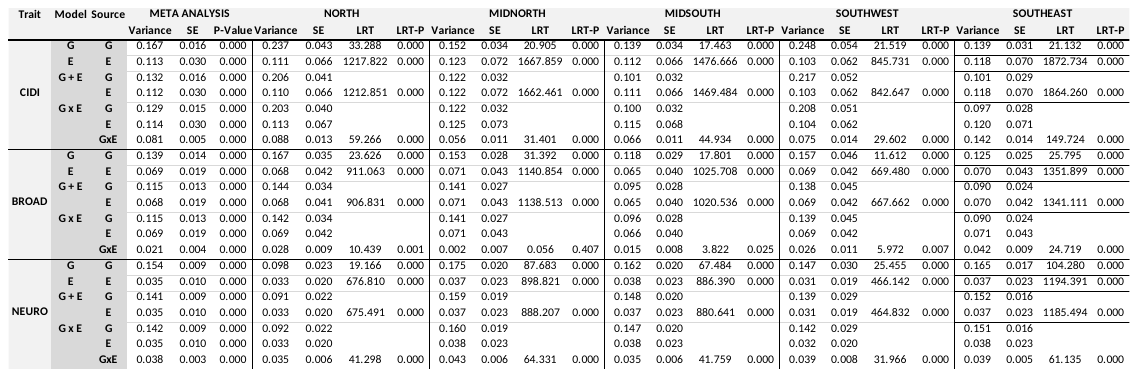

*Abbreviations.* E, environmental relationship matrix using full trauma principal components; SE, standard error; LRT, log-ratio test value; LRT-P, log-ratio test P-Value; G, genetic (Genomic Relationship Matrix); E, environmental (full trauma exposure ERM); GxE, genome-by-trauma exposure interaction.

**Table 13.** Mixed Linear Model Results Including Es of Full Catastrophic Trauma Exposure Principal Components.

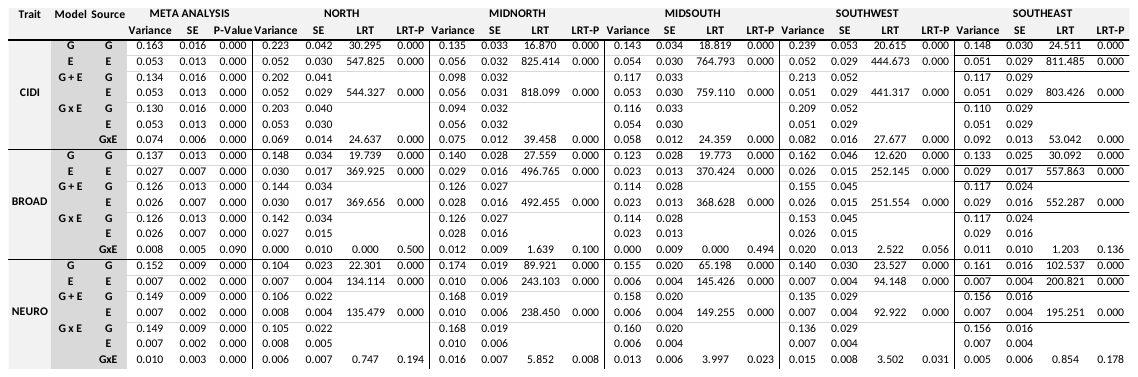

*Abbreviations.* E, environmental relationship matrix using full trauma principal components; SE, standard error; LRT, log-ratio test value; LRT-P, log-ratio test P-Value; G, genetic (Genomic Relationship Matrix); E, environmental (full trauma exposure ERM); GxE, genome-by-trauma exposure interaction.

**Table 14.** Mixed Linear Model Results Including Es of Full Trauma Exposure Principal Components using Unrelated Individuals.

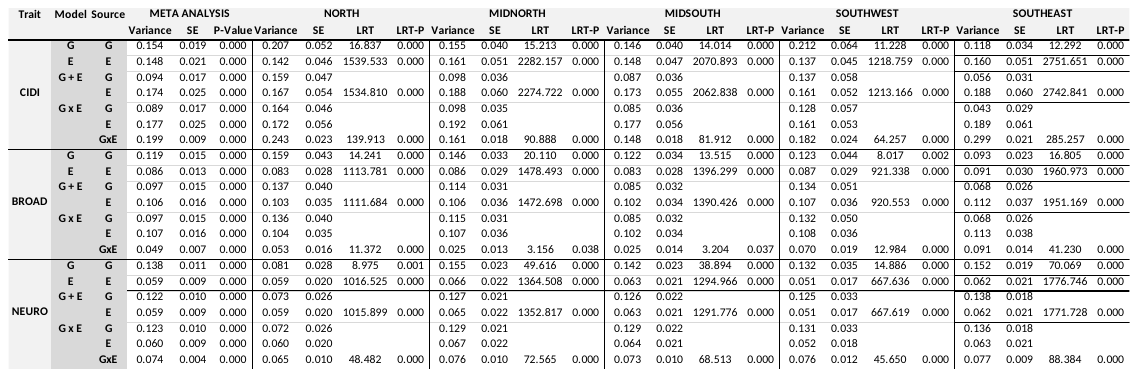

*Abbreviations.* ERM, environmental relationship matrix using full trauma principal components; SE, standard error; LRT, log-ratio test value; LRT-P, log-ratio test P-Value; G, genetic (Genomic Relationship Matrix); E, environmental (full trauma exposure ERM); GxE, genome-by-trauma exposure interaction.

**Table 15.** Mixed Linear Model Results Including Es of Full Trauma Exposure Principal Components Pre-Corrected for Genomic Relationship Matrix.

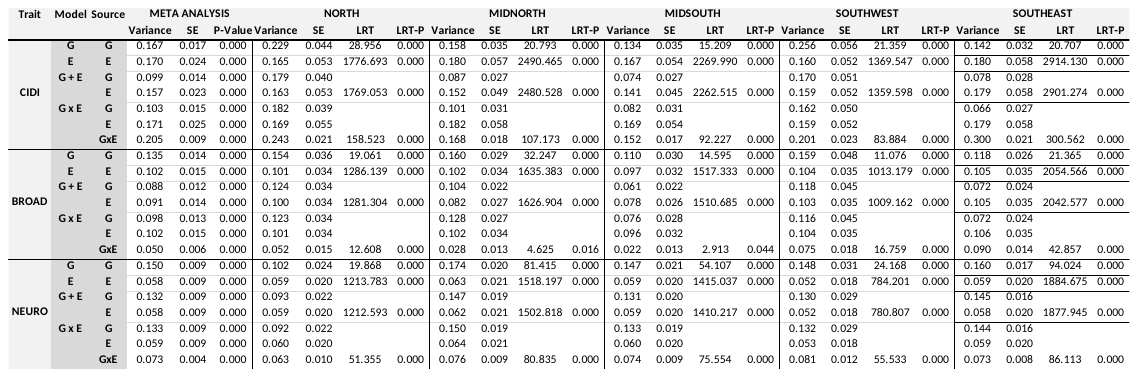

*Abbreviations.* E, environmental relationship matrix using full trauma principal components; SE, standard error; LRT, log-ratio test value; LRT-P, log-ratio test P-Value; G, genetic (Genomic Relationship Matrix); E, environmental (full trauma exposure ERM); GxE, genome-by-trauma exposure interaction.

**Table 16.** Mixed Linear Model Results Including Es of Full Trauma Exposure Principal Component 1.

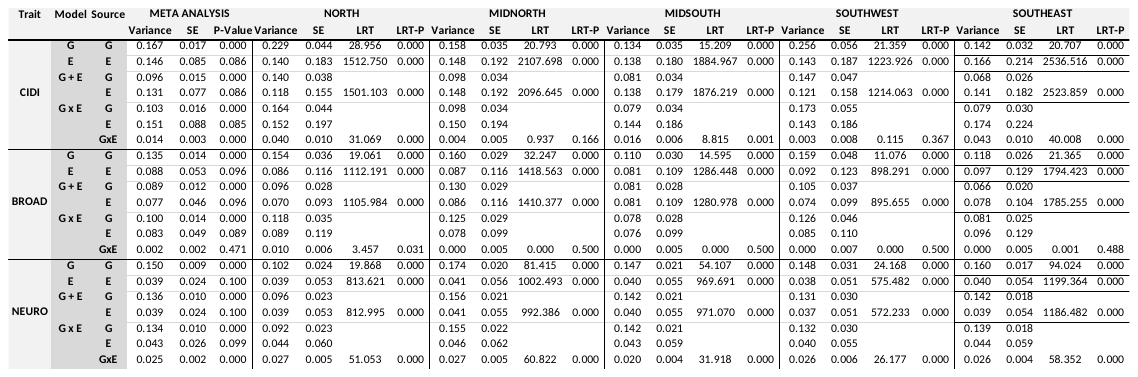

*Abbreviations.* E, environmental relationship matrix using full trauma principal components; SE, standard error; LRT, log-ratio test value; LRT-P, log-ratio test P-Value; G, genetic (Genomic Relationship Matrix); E, environmental (full trauma exposure ERM); GxE, genome-by-trauma exposure interaction.

**Table 17.** Mixed Linear Model Results Including Es of Female Only Full Trauma Exposure Principal Components.

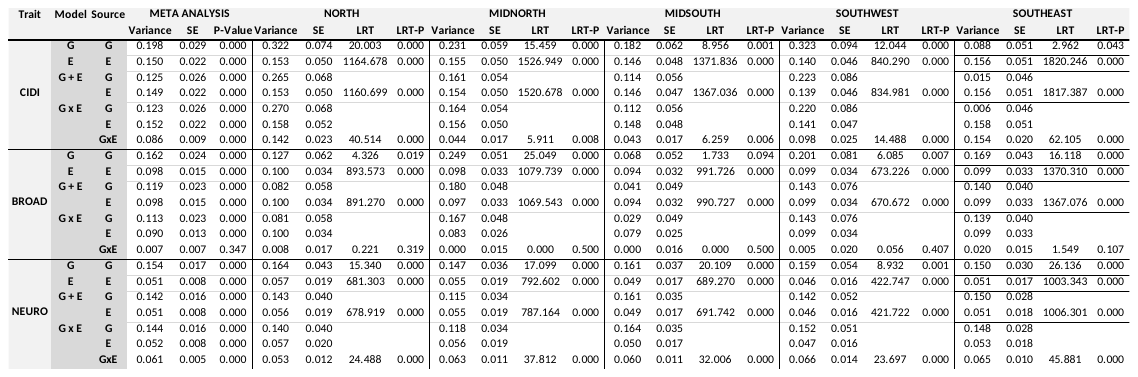

*Abbreviations.* E, environmental relationship matrix using full trauma principal components; SE, standard error; LRT, log-ratio test value; LRT-P, log-ratio test P-Value; G, genetic (Genomic Relationship Matrix); E, environmental (full trauma exposure ERM); GxE, genome-by-trauma exposure interaction.

**Table 18.** Mixed Linear Model Results Including Es of Male Only Full Trauma Exposure Principal Components.

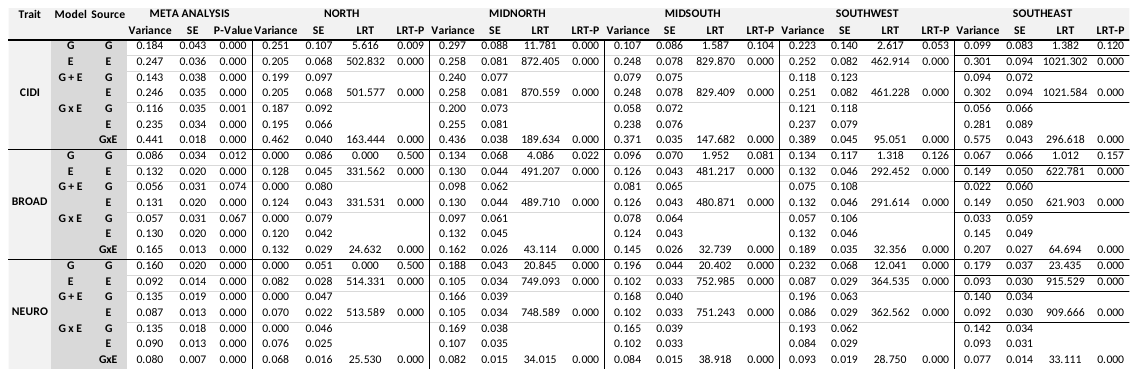

*Abbreviations.* E, environmental relationship matrix using full trauma principal components; SE, standard error; LRT, log-ratio test value; LRT-P, log-ratio test P-Value; G, genetic (Genomic Relationship Matrix); E, environmental (full trauma exposure ERM); GxE, genome-by-trauma exposure interaction.

**Table 19.** Mixed Linear Model Results Including Es of Female Only Full Trauma Exposure Principal Components using Unrelated Individuals.

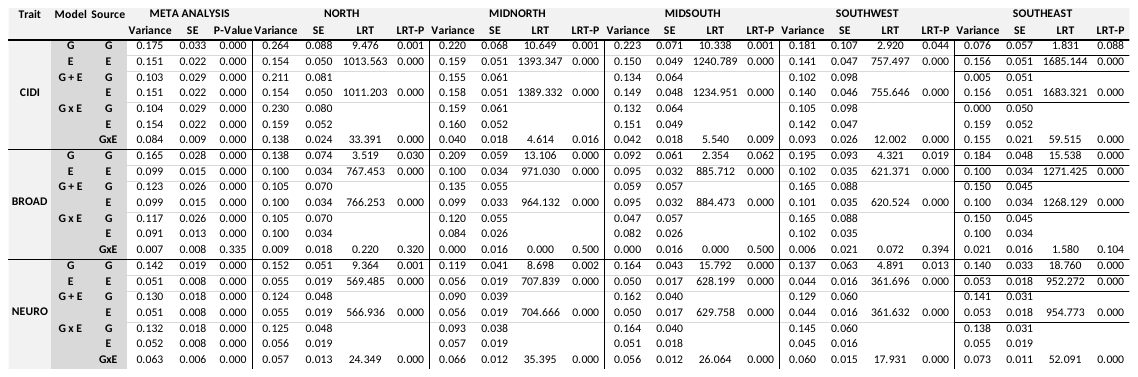

*Abbreviations.* E, environmental relationship matrix using full trauma principal components; SE, standard error; LRT, log-ratio test value; LRT-P, log-ratio test P-Value; G, genetic (Genomic Relationship Matrix); E, environmental (full trauma exposure ERM); GxE, genome-by-trauma exposure interaction.

**Table 20.** Mixed Linear Model Results Including Es of Male Only Trauma Exposure Principal Components using Unrelated Individuals.

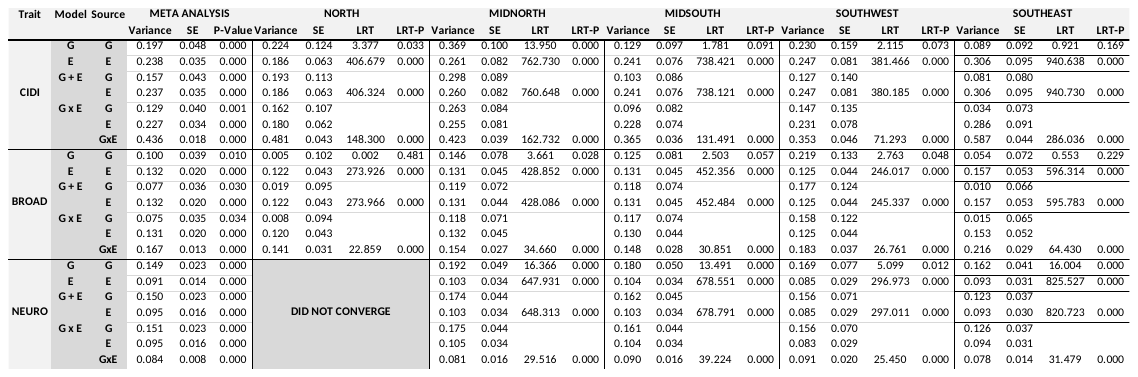

*Abbreviations.* E, environmental relationship matrix using full trauma principal components; SE, standard error; LRT, log-ratio test value; LRT-P, log-ratio test P-Value; G, genetic (Genomic Relationship Matrix); E, environmental (full trauma exposure ERM); GxE, genome-by-trauma exposure interaction.

**Section B**

A consistent pattern observed in results were large LRT values when including environmental relationship matrices (Es) into the mixed linear models (see **Table SB1**). This suggests a substantial improvement in model fit when including the ERMs. Whilst the estimates and standard errors observed for the proportion of variance explained by the Es suggest statistical significance, they do not suggest very *strong* significance, which is discrepant with what the LRT values suggest.

**Table SB1.** North Cluster Mixed Linear Model Results Including E of Full Trauma Exposure Principal Components.

|  |  |  | **NORTH CLUSTER** | |  |  |
| --- | --- | --- | --- | --- | --- | --- |
| **Trait** | **Model** | **Source** | **Variance** | **SE** | **LRT** | **LRT-P** |
| **CIDI** | **G** | **G** | 0.229 | 0.044 | 28.956 | 0.000 |
|  | **E** | **E** | 0.170 | 0.055 | 1801.636 | 0.000 |
|  | **G + E** | **G** | 0.171 | 0.040 |  |  |
|  |  | **E** | 0.169 | 0.055 | 1792.306 | 0.000 |
|  | **G x E** | **G** | 0.174 | 0.039 |  |  |
|  |  | **E** | 0.176 | 0.057 |  |  |
|  |  | **GxE** | 0.241 | 0.021 | 159.722 | 0.000 |

*Abbreviations.* CIDI, Composite International Diagnostic Interview Depression; G, Genomic Relationship Matrix; E, Environmental Relationship Matrix; GxE, Genome-by-Trauma Exposure Relationship Matrix; LRT, Log-Likelihood Ratio Test; P, P-value.

To explore this discrepancy the log-likelihood distribution was examined. A log-likelihood function aims to identify and fit the most appropriate distribution to the data available (e.g. a normal distribution). A value from the observed data will be tested as the mean of the explored distribution. A log-likelihood value, signifying the likelihood of observing the data is obtained. A range of values are tested, and the resulting log-likelihood values are plotted to form the log-likelihood distribution. The highest value i.e. the maximum likelihood estimate, signifies the optimal position of the distribution explored to the data at hand.

Here, the discrepancy suggests a potentially unusual log-likelihood distribution which may be due to the nature of the Es used. Large LRT values would suggest the distribution to have a steep increase, whereas, the smaller standard errors would suggest a plateau surrounding the maximum estimate of the log-likelihood. To plot the log-likelihood distribution, a range of E values and their corresponding log-likelihood value was plotted (**Figure 3**).

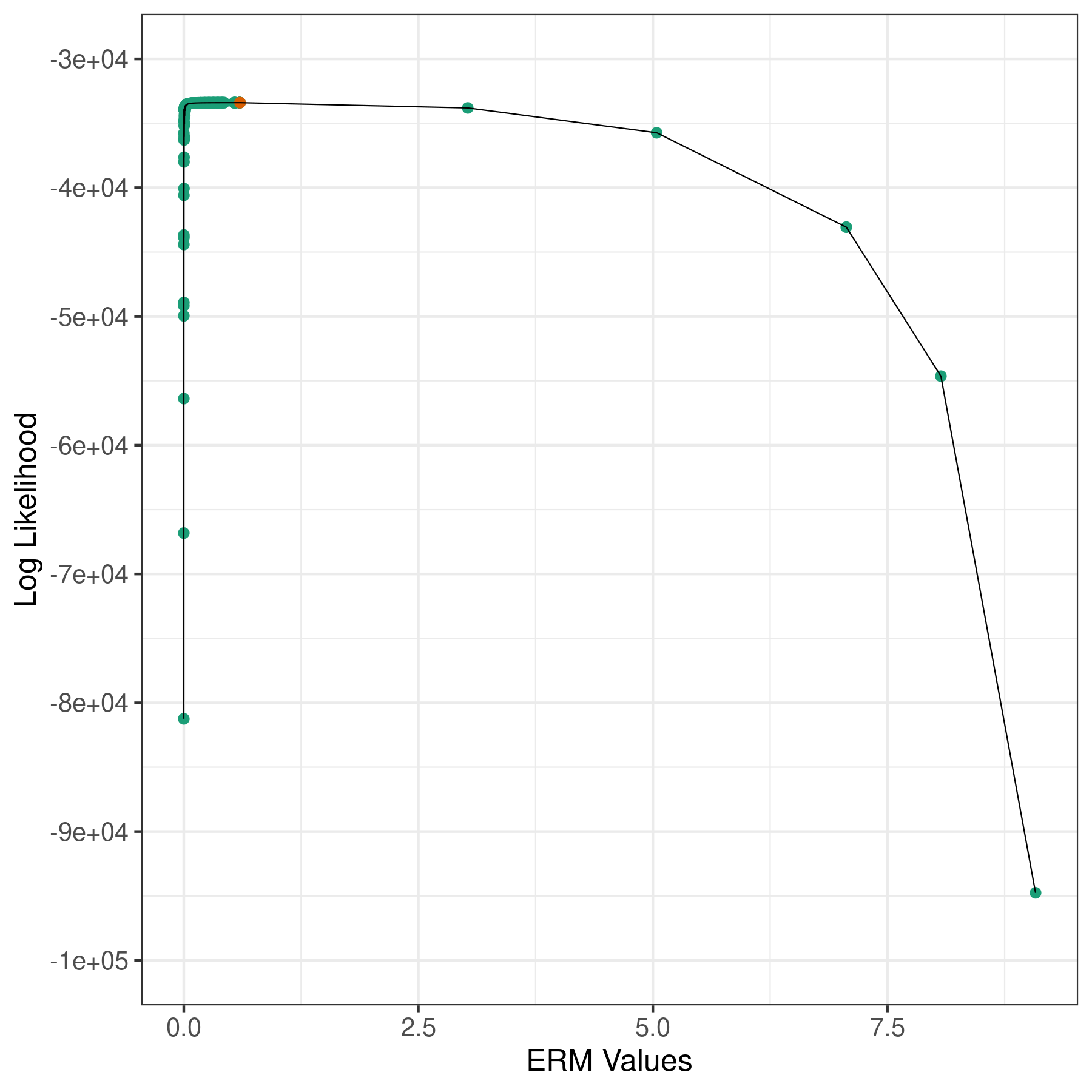

***Figure SB1*.** Log-Likelihood Distribution of Mixed Linear Model Including the Trauma Exposure Environmental Relationship Matrix (ERM) for the UK Biobank North Geographical Cluster and Neuroticism Phenotype. The x-axis represents values of the ERM. The y-axis represents log-likelihood values. The orange point is the maximum likelihood estimate.

The distribution observed explains the discrepancy observed between the LRT values and the estimates/standard errors of the variance attributable to the E.
